# Quantitative proteomics and phosphoproteomics of urinary extracellular vesicles define diagnostic biosignatures for Parkinson’s Disease

**DOI:** 10.1101/2022.01.18.22269096

**Authors:** Marco Hadisurya, Li Li, Kananart Kuwaranancharoen, Xiaofeng Wu, Zheng-Chi Lee, Roy N. Alcalay, Shalini Padmanabhan, W. Andy Tao, Anton Iliuk

**Affiliations:** Tymora Analytical Operations, West Lafayette, IN 47906; Department of Biochemistry, Purdue University, West Lafayette, IN 47907; School of Electrical and Computer Engineering, Purdue University, West Lafayette, IN 47907; Department of Chemistry, Purdue University, West Lafayette, IN 47907; West Lafayette Junior/Senior Highschool, West Lafayette, IN 47906; Department of Neurology, Columbia University Irving Medical Center, New York, NY 10032; The Michael J. Fox Foundation for Parkinson’s Research, New York City, NY 10163; Department of Medicinal Chemistry and Molecular Pharmacology, Purdue University, West Lafayette, IN 47907; Purdue Center for Cancer Research, Purdue University, West Lafayette, IN 4790

## Abstract

Mutations in the leucine-rich repeat kinase 2 (LRRK2) gene have been recognized as genetic risk factors for both familial and sporadic forms of Parkinson’s disease (PD). However, compared to cancer, overall lower genetic mutations contribute to the cause of PD, propelling the search for protein biomarkers for early detection of the disease. Utilizing 141 urine samples from four groups, healthy individuals (control), healthy individuals with G2019S mutation in the *LRRK2* gene (non-manifesting carrier/NMC), PD individuals without G2019S mutation (idiopathic PD/iPD), and PD individuals with G2019S mutation (LRRK2 PD), we applied a proteomics strategy to determine potential diagnostic biomarkers for PD from urinary extracellular vesicles (EVs). After efficient isolation of urinary EVs through chemical affinity followed by mass spectrometric analyses of EV peptides and enriched phosphopeptides, we identified and quantified 4,480 unique proteins and 2,682 unique phosphoproteins. We detected multiple proteins and phosphoproteins elevated in PD EVs that are known to be involved in important PD pathways, in particular the autophagy pathway, as well as neuronal cell death, neuroinflammation, and formation of amyloid fibrils. Our data revealed that LRRK2 and its Rab substrates are altered but not significant PD biomarkers. We established two panels of proteins and phosphoproteins as novel candidates for disease and risk biomarkers, and substantiated using ROC, machine learning, clinical correlation, and in-depth network analysis. Several disease biomarkers were further validated in patients with PD using parallel reaction monitoring (PRM) and immunoassay for targeted quantitation. These findings demonstrate a general strategy of utilizing biofluid EV proteome/phosphoproteome as an outstanding and non-invasive source for a wide range of disease exploration.

## Introduction

It has been more than two centuries since Parkinson’s disease (PD) was described by Dr. Parkinson in 1817^1^. PD is the second most common neurogenerative disorder after Alzheimer’s disease (AD)^2^. PD’s most common pathological finding is a decreased pigmentation in the substantia nigra pars compacta (SNpc) caused by the death of dopaminergic neurons, leading to progressive deterioration of motor function^3, 4^. In addition to motor symptoms, non-motor symptoms may include cognitive impairment, autonomic dysfunction, hyposmia, and sleep disturbances^5^. Currently, PD is incurable and progresses gradually with symptom deterioration into severe disabilities^6^. It has been estimated that PD affects 1 percent of the population over 60^7^. Overall, as many as 1 million Americans are living with PD, and approximately 60,000 Americans are diagnosed with PD each year^8, 9^.

While the cause of PD is currently unknown, researchers speculate that environmental and genetic factors contribute to its development^10^. Large-scale genome-wide association studies (GWAS) have identified 41 independent risk variants for PD in various cohorts^4, 11^. A subset of patients develops PD because of a major genetic risk. Specifically, mutations in the Leucine-rich repeat kinase 2 (*LRRK2*) gene are found in hereditary forms, emphasizing the shared molecular pathway driving both familial and non-familial PD to comprise the most common cause of the disease^12, 13^. Mutations in *LRRK2* have been recognized as genetic risk factors for sporadic (∼1%) and familial forms of PD (∼5%)^13^. *LRRK2* encodes a large protein of 2,527 amino acids containing two functional enzymatic domains, the GTPase and the Ser/Thr kinase domains, and several protein-protein interaction domains such as the armadillo, ankyrin, leucine-rich repeat (LRR), and WD40 domains^14, 15^. Out of many mutations in *LRRK2*, Gly2019→Ser (G2019S) mutation in its kinase domain is by far the most common among caucasians^16^. Interestingly, some individuals with the G2019S mutation, known as the non-manifesting carrier (NMC) group, do not develop PD. Whether they will develop the disease at an older age remains unclear.

Recent findings regarding the Gly2019→Ser (G2019S) mutation in the *LRRK2* kinase domain have uncovered that the mutation drives changes in vesicular trafficking, autophagy, and lysosomal dysfunction signaling pathways^16^. The changes in these signaling pathways are attributed to the hyperactivation of the LRRK2 kinase activity assessed by phosphorylation of its substrates, the Rab proteins^17^. Rab proteins are the main regulators of important aspects of autophagy and lysosome activity, including membrane trafficking, vesicle formation, vesicle movement along actin and tubulin networks, and membrane docking and fusion. In short, from the evidence above, it is conceivable that the changes in signaling pathways caused by the Gly2019→Ser (G2019S) mutation in the *LRRK2* kinase domain may be reflected in extracellular vesicles (EVs). Therefore, EVs offer a promising source for protein biomarkers in PD.

EVs (primarily exosomes and microvesicles) are lipid bilayer-coated nanoparticles secreted by all cell types. The secretion of EVs was initially considered a means of eliminating proteins, lipids, and RNA from inside the cells^18^. With accumulating evidence, EVs have become recognized as a very important component in intercellular communication^19^. Recent studies have reported EVs as a rich resource of biomarkers for the non-invasive detection of neurodegenerative diseases from biofluids^20^. These EV-based disease markers can be identified well before the onset of symptoms or physiological detection of illness, making them promising candidates for early-stage PD diagnosis^21, 22^. Moreover, since phosphorylation events directly reflect cellular physiological status during neurodegeneration, urinary EVs represent a highly promising source of phosphoproteins as non-invasive disease markers^23, 24^. Previous studies from our group have identified numerous EV phosphoproteins in urine and plasma from breast cancer, chronic kidney disease, and kidney cancer patients^25–28^.

Moreover, some other groups already explored the possibility of using EV biofluids, such as cerebrospinal fluid (CSF), saliva, and plasma, as the source for PD diagnosis. CSF exosomes from patients with PD and dementia with Lewy bodies contain a pathogenic species of a-synuclein, which could initiate oligomerization of soluble a-synuclein in target cells and confer disease pathology^29^. Saliva-derived EVs from PD patients have elevated levels of oligomeric α-syn compared to controls^30^. Serum neuronal exosomes of α-synuclein and clusterin could predict and differentiate PD from atypical parkinsonism^31^. Although CSF and plasma-derived EVs have been used for biomarker studies of PD, urine-derived EVs offer another promising clinically viable matrix for PD detection since urine can be non-invasively collected frequently in large volumes and repeatedly at different timepoints^32^. More importantly, although most urine-derived EVs originate from the kidney and urinary tract, a significant proportion of those also originate from distal organs, including the brain^33, 34^. Moreover, urinary EVs will reflect the whole physiological changes that happened to the body of PD patients. Therefore, urine-derived EVs may provide diagnostic opportunities for PD^35, 36^.

Here we present a strategy for the discovery and development of proteins and phosphoproteins from urinary EVs as diagnostic biosignatures for Parkinson’s disease. For the discovery experiment, we utilized 82 individual urine samples made available from Columbia University Irving Medical Center (hereinafter referred to as “Columbia LRRK2 cohort”) under a Michael J. Fox Foundation (MJFF)-funded LRRK2 biomarker project^32^ and split them into 164 analyses (82 proteomics and 82 phosphoproteomics). We used our in-house developed unique EVtrap (<u>E</u>xtracellular <u>V</u>esicles total recovery <u>a</u>nd <u>p</u>urification) approach to efficiently enrich EVs and coupled it with LC-MS-based detection and quantitation for accurate urinary EV proteome and phosphoproteome analysis^37^. EVtrap, based on functionalized magnetic beads with a combination of lipophilic and hydrophilic groups, has a unique affinity toward lipid bilayer membrane coating EVs. EVtrap enables fast and reproducible EV isolation from urine samples. Our approach to date is the first such method to successfully demonstrate the feasibility of developing biofluid-derived EV phosphoproteins for disease profiling^26, 27^. In total, we determined two panels of unique proteins and phosphoproteins as novel high-confidence candidates for disease and risk biomarkers. Disease biomarkers will help diagnose whether a patient currently has PD; on the other hand, risk biomarkers will predict the likelihood of developing PD in the future, especially for those NMC individuals. Our large-scale LC-MS analysis efforts combined with extensive bioinformatics analysis led to the discovery of unique biosignatures for potential Parkinson’s disease diagnostics.

Furthermore, we also analyzed our biomarker involvement in important disease-relevant pathways, which might provide new information for PD intervention. These findings will enhance the discovery and development of novel EV protein-based biomarkers and help create an effective early-stage clinical diagnosis strategy for PD. An in-depth understanding of those biosignature pathways could also lead to the potential discovery of new drugs for optimal intervention strategies in PD progression.

## Results

### Urine EV phosphoproteomics study design and sample quality control for PD biosignature development

For decades, scientists have been focusing on PD genotype marker discovery. In the case of *LRRK2* G2019S mutation diagnosis alone, people are often found to either have the mutation in their genome without even experiencing PD (non-manifesting carrier/NMC) or do not have the mutation in their genome although they are suffering from PD (idiopathic PD/iPD). Furthermore, whether people with diagnosed NMC will develop PD later in their lives remains unclear. Multiple recent studies have shown that analysis of proteins and phosphoproteins in many cases provides a better snapshot of cellular processes and disease progression than genomic or transcriptomic investigations^38–41^. Proteome/phosphoproteome profiling efforts have already demonstrated significant advantages for disease diagnosis and prediction of treatment response^42–45^. This is particularly true for kinase-dependent conditions and kinase inhibitor drugs^46–48^. Using this study design, we have further confirmed what is already known in the PD research community - that genotype markers are unreliable. Therefore, there is a critical need to shift the focus to developing protein- and phosphoprotein-based biomarkers for PD detection instead. Since the *LRRK2* G2019S mutation alters the phosphorylation activity and these changes are reflected in extracellular vesicles, this supports the rationale behind using EVs as promising biosignature sources for PD diagnosis. Moreover, considering that many phosphorylation events directly reflect cellular physiological status, urinary EVs represent a highly promising source of proteins and phosphoproteins as non-invasive PD markers^23, 24^. This is further reinforced by the recent studies showing Parkinson’s disease relevance of LRRK2 phosphorylation in urinary EVs^49–51^ and *LRRK2* G2019S mutation influence on neat urine proteome^52^.

Urine samples were collected at Columbia University Irving Medical Center (CUIMC) from four cohorts with or without PD, and with or without the common G2019S mutation in the *LRRK2* gene^32^. The samples were collected from March 2016 to April 2017 under a Michael J. Fox Foundation (MJFF)-funded LRRK2 biomarker project. The participants underwent clinical evaluation of their cognitive functions using the Montreal Cognitive Assessment (MoCA) and motor skills using the Unified Parkinson’s Disease Rating Scale part III (UPDRS-III). This sample cohort has been uniquely curated for in-depth analysis and comparison of *LRRK2* genotype and activity effects on PD as previously described^32, 53^. These 141 samples were divided into three groups: the discovery experiment (82 samples) and two validation experiments (56 samples) (**Figure 1**). We were fully blinded to the identity of all samples until after the complete analysis. These four groups - control, NMC, iPD, and LRRK2 PD - were the major components of this biosignature study design. The demographic information for all samples is provided in **Table 1**. As shown, the four groups were balanced for all demographic variables.

**Figure 1.**
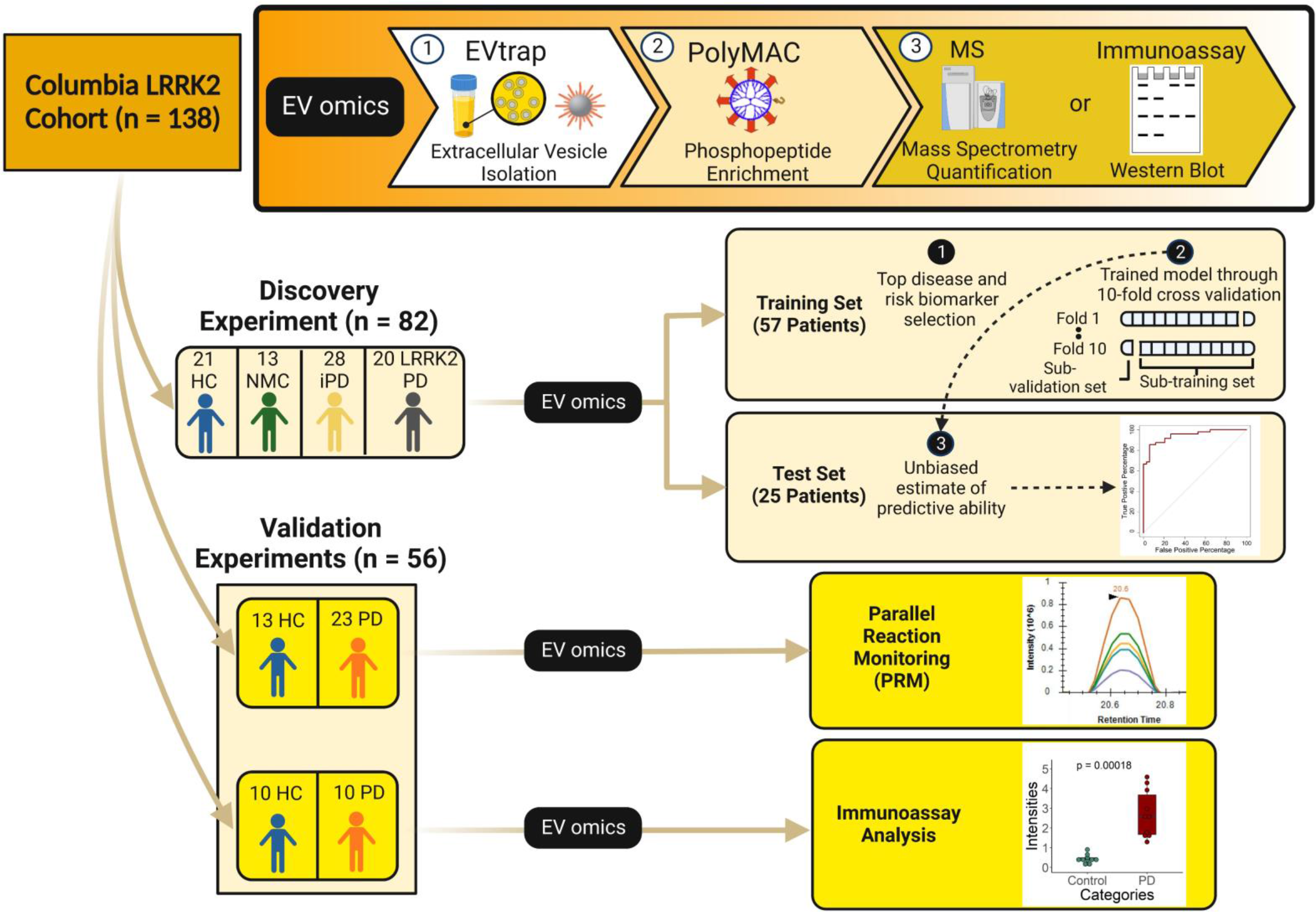
The development and validation of biomarker signatures for the diagnosis of Parkinson’s Disease. A total of 141 urine samples were divided into two groups: the discovery and validation experiments. The urine samples were processed using our in-house ①EVtrap for EV isolation and ②PolyMAC (where applicable) for phosphopeptides enrichment. In the discovery experiment, the available 82 clinical urine samples were further randomly distributed into training and test sets for biomarker prediction. We proposed categorizing the potential biomarkers in 2 main categories: disease markers as the potential biomarkers for PD regardless of the *LRRK2*-G2019S mutation and risk markers as the potential biomarkers for the risk of acquiring PD. Utilizing machine learning, we discovered the top disease and risk biomarkers. Furthermore, we also trained our model using the ten-fold cross-validation and unbiasedly estimated the predictive ability on the test set. For biomarker validation, another 56 clinical urine samples were divided into two groups for parallel reaction monitoring (PRM) and immunoassay analysis. HC: healthy control, NMC: non-manifesting carrier, iPD: idiopathic Parkinson’s Disease, LRRK2 PD: LRRK2 Parkinson’s Disease, and PD: Parkinson’s Disease.

**Table 1.** The summary of cohort demographics and clinical characteristics for all 82 patients whose samples were used in the discovery experiment. The range units for age and disease duration are in years (see **Supplementary Data 12** for more details). The groups include healthy individuals (control), healthy individuals with G2019S mutation in the *LRRK2* gene (non-manifesting carrier/NMC), PD individuals without G2019S mutation (idiopathic PD/iPD), and PD individuals with G2019S mutation (LRRK2 PD).

| Demographics | Control | NMC | iPD | LRRK2 PD | Overall |
| --- | --- | --- | --- | --- | --- |
| Age (range; years) | 69.4 (59-85) | 58.5 (37-83) | 66.1 (45-82) | 69.7 (56-90) | 66.6 (37-90) |
| Gender<br>(Female/Male) | 10/11 | 6/7 | 3/4 | 9/11 | 37/45 |
| Disease Duration<br>(range; years) | n/a | n/a | 0-18 | 1-26 | 0-26 |
| MoCA | 27.7 (24-30) | 28.5 (27-30) | 27.2 (23-30) | 27.2 (22-30) | 27.5 (22-30) |
| UPDRS-III | 1.1 (0-5) | 0.8 (0-3) | 17.5 (5-38) | 21.1 (1-53) | 11.5 (0-53) |

To evaluate the quality of samples and demonstrate the superior efficiency of isolating urinary EVs by EVtrap, we first selected a few representative samples and analyzed them using Tunable Resistive Pulse Sensing (TRPS), Western blotting with anti-CD9 and anti-LRRK2 antibodies, and LC-MS analyses. Nanoparticle size and distribution analysis with qNano (TRPS) of EVtrap- and ultracentrifugation (UC)-enriched urine EV samples both demonstrated a similar range of the isolated EVs, with the majority being in the 100-200 nm range (**Supplementary Figs. 1a, b**). Here, EVtrap showed a higher concentration of isolated EVs, as demonstrated in a previous publication^37^. Similarly, detection of CD9 and LRRK2 target proteins by Western blot from 5 randomly selected urine samples revealed a significant increase in signal levels for both proteins after EVtrap isolation compared to UC (**Supplementary Fig. 1c**). Finally, to show the reproducibility of our analytical procedure (from EVtrap enrichment to LC-MS analysis), we split a human urine sample into six aliquots and processed them separately for LC-MS analysis. The raw intensities were log-based 2 transformed, filtered (70% quantification), and imputed before the coefficient of variation (CV) calculation. **Supplementary Fig. 2a** demonstrates outstanding reproducibility of the procedure, with almost all of the proteins detected and quantified across all six samples falling under 10% CV) and the vast majority under 5% CV (complete proteomics data in **Supplementary Data 1**). Moreover, we identified 90 EV markers from the top 100 EV markers and common EV proteins as listed in ExoCarta, such as CD9, CD63, and CD81, which were included as minimal information for studies of extracellular vesicles 2018 (MISEV2018)^54, 55^. MISEV 2018 guidelines also proposed that it is more appropriate to show depletion than to expect a binary presence/absence of proposed urine negative markers in urinary EVs. To show the depletion of high-abundant free urine proteins after EVs isolation, the raw intensities for all quantified proteins in 12 samples (6 pairs of direct urine and urinary EVs isolated using EVtrap, where each pair was from the same urine sample with the same volume) were log-based 2 transformed, filtered (6 minimal valid values in at least 1 group), and imputed before the fold change calculation. As expected, we showed in **Supplementary Fig. 2b** that some of the most abundant proteins in urine, such as ALB (Fold-change = −181), UMOD (Fold-change = −65), and AMBP (Fold-change = −596), were significantly depleted in urinary EVs (paired Student’s two-tailed t-test p-values were shown, see **Supplementary Data 1** for complete data)^56^.

### Urinary EVs as prominent sources of PD biomarkers

We processed 82 urine samples individually for the discovery experiment following the illustrated workflow in **Supplementary Fig. 2c** using approximately 10-15mL of each urine after normalization by creatinine concentration. As the first step, we employed EVtrap to capture the complete EV profile from the urine samples using the synthesized magnetic beads described previously^37^. Following EV elution and drying, we lysed them with the optimized phase-transfer surfactant-based procedure to extract and denature proteins. After the reduction/alkylation step, the proteins were digested with sequential Lys-C and trypsin additions, and the resulting peptides were desalted. Here, a small portion of each sample (∼1%) was used for direct proteomic analysis. We carried out phosphopeptide enrichment using our in-house developed dendrimer-based PolyMAC method on the remaining majority of each sample and analyzed by LC-MS. Indexed Retention Time Standard containing 11 artificial synthetic peptides was added to all proteomic and phosphoproteomic samples for improved peptide quantitation and reproducibility. The samples were analyzed by Thermo Fisher Q-Exactive HF-X MS coupled with the Ultimate 3000 UHPLC system.

Our urinary EV proteomic and phosphoproteomic analyses identified and quantitated 4,480 unique proteins from 46,240 peptide groups and 2,682 unique phosphoproteins from 10,620 phosphopeptide groups (**Supplementary Data 2** and **3;** see **Fig. 2a** for quantified features). We evaluated whether our identified EV proteins and phosphoproteins were a good source for PD assessment. We compared our data with the brain-specific RNA-seq data downloaded from the Human Protein Atlas website^57^. We used 2,587 proteins classified as brain-elevated from the Human Protein Atlas dataset to compare our EV protein and phosphoprotein data. We found that 8.9% of our EV proteins were denoted as brain-elevated (**Supplementary Fig. 3**). While the brain is likely a minimal source of EV proteins in urine^58^, this finding strengthens our hypothesis that urinary EV proteins and phosphoproteins are great candidates as potential biomarkers for PD.

**Figure 2.**
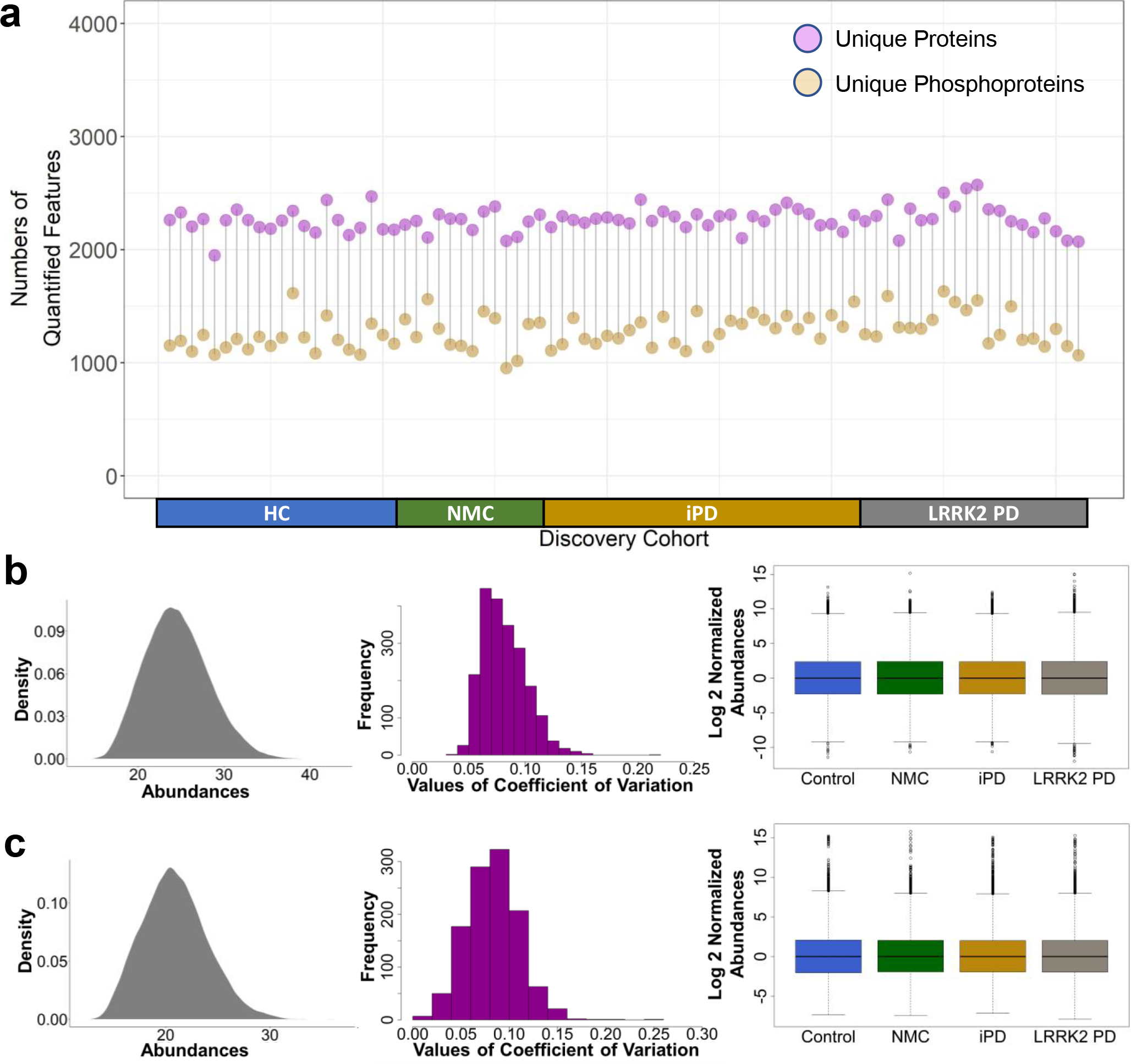
The summary of identification and quantification for all 82 patients. a) Cleveland Dot Plots for all quantified proteins and phosphoproteins. Both proteomic (b) and phosphoproteomic data (c) were normalized based on internal standards. Most quantified proteins and phosphoproteins had a CV of less than 20%. Normalized quantified data in the training set were then analyzed using feature selection to find potential biomarkers for PD.

The proteins and phosphoproteins identified must pass a rigorous statistical threshold and normalization to be statistically useful, as explained in more detail in the materials and methods section. We normalized both the proteome and phosphoproteome data based on internal standards. **Figures 2b** and **2c** confirmed that the data had been effectively normalized with a coefficient of variation (CV) less than 20%. Before we divided the discovery experiment data into the training and test sets, we performed gene ontology, clinical parameter correlation, and pathway analyses using the complete data (n = 82).

### Functional annotation identifies immune response, complement activation, and vesicle-mediated transport as the most prominent etiologies of PD in urine EVs

We identified the upregulated proteins and phosphoproteins in NMC, iPD, and LRRK2 PD groups against the controls (permutation-based FDR < 0.05 and log base 2 fold-change > 0.5, which equals to ∼1.414 fold-change, see **Supplementary Data 4** for the complete proteomic data and **Supplementary Data 5** for the complete phosphoproteomic data). Focusing on the upregulated proteins, we performed gene ontology analysis to understand the correlation between all upregulated proteins and PD. We utilized STRING database (v11.5) for biological process gene ontology analysis^59^. The gene ontology analyses were set with a threshold FDR of 5% after Benjamin-Hochberg correction. Select gene ontology results are shown in **Supplementary Fig. 4** and listed in **Supplementary Data 6**.

The abnormal glycation and glycosylation seem to be more common than previously thought in PD and may underlie inflammation and mitochondria-induced oxidative stress in a feed-forward mechanism^60^. Furthermore, since PD patients’ CSF appears to have a specific metabolomic signature that reflects alterations in glycation or glycosylation, it was not surprising to discover that some biological process alterations involving glycosylation were also enriched the patients’ in urine EVs^60, 61^. Interestingly, we discovered that glycoside metabolic process was upregulated in NMC (**Supplementary Fig. 4a**). As seen in **Supplementary Fig. 4b**, cytolysis, in which various reducing agents, including dopamine, inhibit, was also enriched in iPD^62^. The fact that dopamine production is diminished in PD supports the observed increase in cytolysis. In addition, the immune response and the complement activation, which is a part of innate immune system, are both enhanced in iPD. Complement activation, a major inflammatory mechanism in PD, on melanized neurons tends to increase in the PD substantia nigra^63^. When investigated using magnetic resonance imaging, the signal intensity of melanized neurons in the substantia nigra pars compacta in PD patients was greatly reduced, suggesting that the increase of complement activation contributes to PD development^64^. It has been hypothesized that aging-related metabolic changes could contribute to the progression and onset of PD^65^. Therefore, it is not surprising to see that cellular catabolic and carbohydrate derivative catabolic processes were upregulated in LRRK2 PD (**Supplementary Fig. 4c**). Moreover, since LRRK2 substrates are involved in membrane trafficking, vesicle-mediated transport was also found to be enhanced in LRRK2 PD.

**Supplementary Fig. 4d** comparison provides an interesting discovery about the difference between iPD and LRRK2 PD gene ontology. In this evaluation, cell adhesion molecule binding, lysosome, leukocyte transendothelial migration, and adaptive immune system were enhanced in LRRK2 PD. Lysosome activity was upregulated due to LRRK2 substrates’ involvement in lysosome sorting, degradation, and autophagy^66^. Leukocyte transendothelial migration, which is crucial for innate immunity and inflammation, and the adaptive immune system, which is carried out by lymphocytes, were enriched, indicating the potential that *LRRK2* G2019S mutation could further amplify the already pro-inflammatory function of LRRK2 in inflammasome activity^67^. Although the expression of LRRK2 is mainly thought of in the context of neurons, it is also discovered to be highly expressed by immune cells such as monocytes, macrophages, and B cells, where LRRK2 direct substrate mediated vesicle trafficking is heavily involved in their immune response initiation^67, 68^.

### Correlation of proteome and phosphoproteome profiles with clinical parameters

We investigated any correlations between the expression of the proteins and phosphoproteins with age, gender, disease duration, MoCA score, and UPDRS-III score of the patients (see **Supplementary Data 7** for the cohort demographics and clinical characteristics). There is increasing evidence that sex is an important factor in the development of PD^69^. In men, the risk of developing PD is nearly twice as high as in women. However, women have a higher mortality rate and faster disease progression^70^. MoCA was initially designed to evaluate mild cognitive impairment associated with AD to assess memory, executive functions, and verbal fluency, among others, and can be applied in a short period of time^71^. The test has been used for the cognitive evaluation of patients with PD to identify cognitive deficits. MoCA scores range between 0 and 30, where a score of 26 or over is considered normal. UPDRS-III scoring method evaluates the patient’s motor skills ranging from 0 to 108, with 108 being the worst.

We found that the expression levels of ENPEP, GDPD3, NAGA, NEDD4L, QPRT, and SCAMP3 proteins in urine EVs were significantly higher (p-value < 0.05, calculated using the unpaired two-samples Wilcoxon test) in males than in females (**Supplementary Fig. 5a**). There were positive correlations in the expression of FUT6 (R^2^=0.83, P<0.05) and HAO2 (R^2^=0.90, P<0.005) proteins with age in the female NMC group, as seen in **Supplementary Fig. 5b**. Meanwhile, the expression of ALPL protein was negatively correlated with disease duration in the female iPD group (**Supplementary Fig. 5c**). Related to the MoCA scores in the male NMC group, we found a positive correlation in CAPN5 and HNRNPA1 proteins, and a negative correlation in ENPEP, GDPD3, and GPD1L proteins (**Supplementary Fig. 5d**). Additional significant correlations between protein abundance levels, MoCA scores, and gender are shown in **Supplementary Fig. 5d**.

At the phosphoprotein level, pNEU1 abundance was positively correlated with age in the female NMC group (R^2^=0.86, P<0.01) (**Supplementary Fig. 6a**). DTD1 phosphorylation was positively correlated with MoCA in the female NMC group (**Supplementary Fig. 6b**). pANXA11 and pHLA-B were negatively correlated with MoCA in the male NMC group, while there were positive correlations for CYSRT1, LTB4R, and TJP3 phosphoproteins. In addition, MoCA in female NMC was negatively correlated with the expression of CYSRT1 phosphoprotein. Lastly, the pLTBR4 level in the male LRRK2 PD group was positively correlated with MoCA.

Furthermore, we assessed the correlations of UPDRS-III scores with the protein and phosphoprotein intensities in iPD and LRRK2 PD patients versus those in the healthy individuals. We found several proteins and phosphoproteins depicted in **Figure 3a** and **Supplementary Fig. 7** to be positively and negatively correlated with UPDRS-III, and many that are moderately correlated with the UPDRS-III (0.5 < Pearson correlation < 0.7). Significantly correlated proteins with an FDR of 5% after Benjamin-Hochberg correction and Pearson correlation more than 0.5 are labeled in the volcano plots. Lastly, **Figure 3b** shows three proteins, PEBP4, NEDD4L, and KLK6, with higher than 0.7 Pearson correlation scores, denoting a strong correlation with UPDRS-III. These correlation data need to be further validated, and their relevance to PD evaluated in a translational manner.

**Figure 3.**
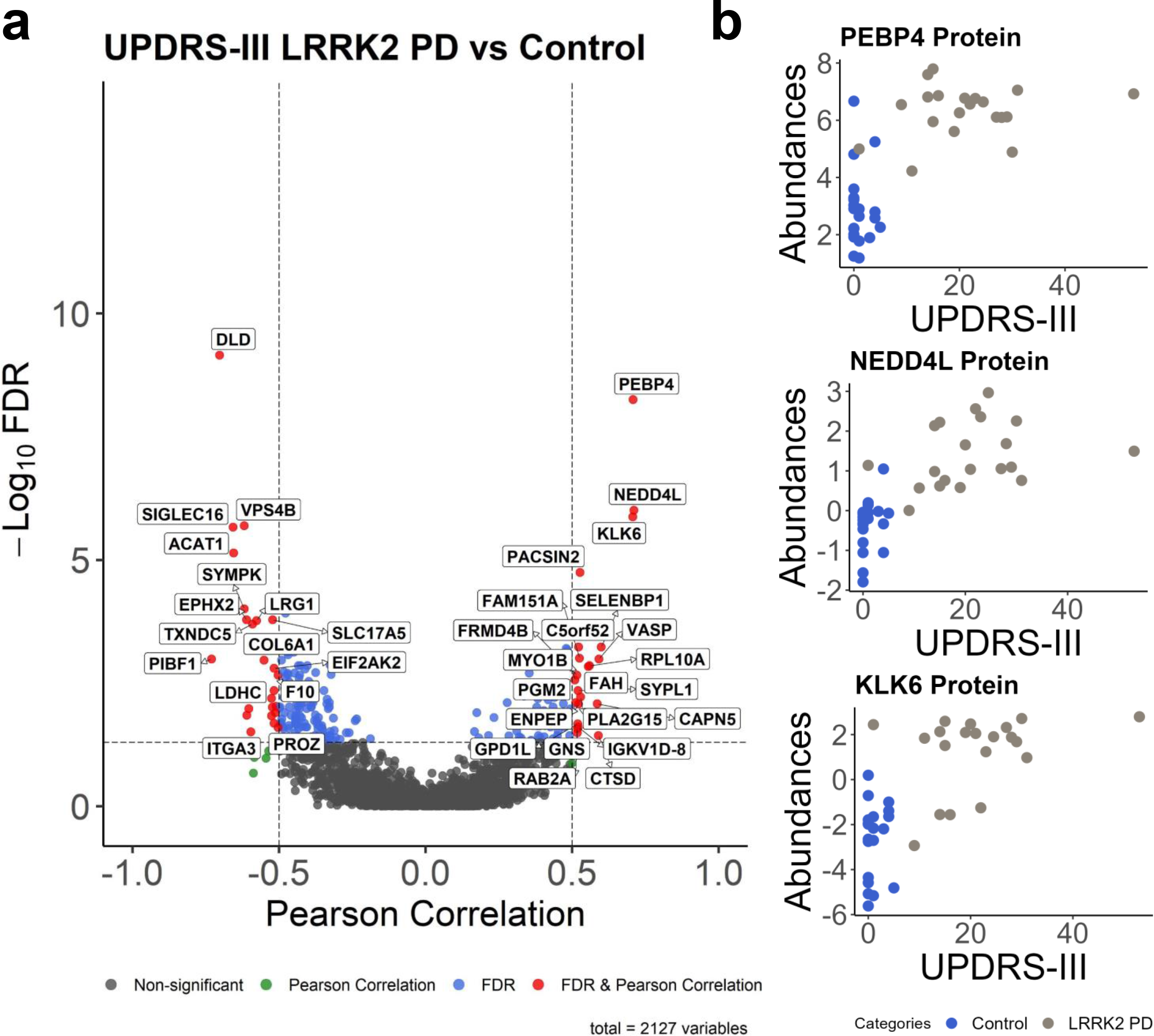
Correlations with clinical parameter, UPDRS-III. a) Pearson correlation scores and associated *P*-values [-log_10_] of all protein intensities with the UPDRS-III score. LRRK2 PD patients were included. Significantly correlated proteins with an FDR of 5% after Benjamin-Hochberg correction are labeled. b) The scatterplots of three biomarkers with strong Pearson correlation scores (> 0.7).

### Disease-related EV protein and phosphoprotein biomarkers are prominently involved in the autophagy pathway

All of the following pathway analyses have been previously reported to be involved in PD onset and progression, but not by the direct experimental evidence of the pathogenic mechanism performed in this study. As potential disease markers, HNRNPA1, IDE, and STK11 proteins are shown to be involved in certain pathways that are important in PD progressions, such as protein targeting to peroxisome, AMPK signaling pathway, leukocyte activation, and mRNA splicing (**Figure 4a**, see **Supplementary Data 8** for protein-protein interactions). These markers also interact closely with PRKACA, VDAC1, VDAC2, and VDAC3, known to be PD related^72^. Moreover, four top disease markers, PCSK1N, HNRNPA1, pPLA2G4A, and pLTB4R, are known to be involved in such important PD pathways as neuronal cell death, neuroinflammation, autophagy, and formation of amyloid fibrils (**Figure 4b**; see **Supplementary Data 9** for more details). From the Ingenuity Pathway Analysis (IPA), the upregulation of IDE leads to neuronal cell death activation, while the upregulation of STK11 indirectly leads to autophagy activation. PLA2G4A and LTB4R phosphoproteins were shown to be involved in downstream GPCRs and MAPK signaling pathways (**Supplementary Fig. 8a**; see **Supplementary Data 10** for table format). Meanwhile, the presence of NEU1, a lysosomal enzyme and a disease marker, supports the emerging concept that PD is a lysosomal disorder^73^ (**Supplementary Fig. 8b**; see **Supplementary Data 10** for table format). Furthermore, the overexpression of PLA2G4A, LTB4R, and NEU phosphoproteins can trigger the autophagy pathway, one of the hallmark pathways in PD (**Supplementary Fig. 8c**; see **Supplementary Data 11** for more details). Interestingly, NEU1 showed two contradicting downstream effects. The overexpression of low-density lipoprotein (LDL)-cholesterol by NEU1 inhibited autophagy. On the other hand, the inhibited expression of high-density lipoprotein (HDL)-cholesterol by NEU1 triggered autophagy. In **Supplementary Fig. 8c**, PLA2G4A is shown to indirectly activate autophagy, supporting the fact that PLA2G4A activation leads to the impairment of autophagy flux by directly increasing lysosomal membrane permeabilization (LMP)^74^. The interactions of LTB4R/RAC1/PAK1/p38 MAPK are also known to activate autophagy.

**Figure 4.**
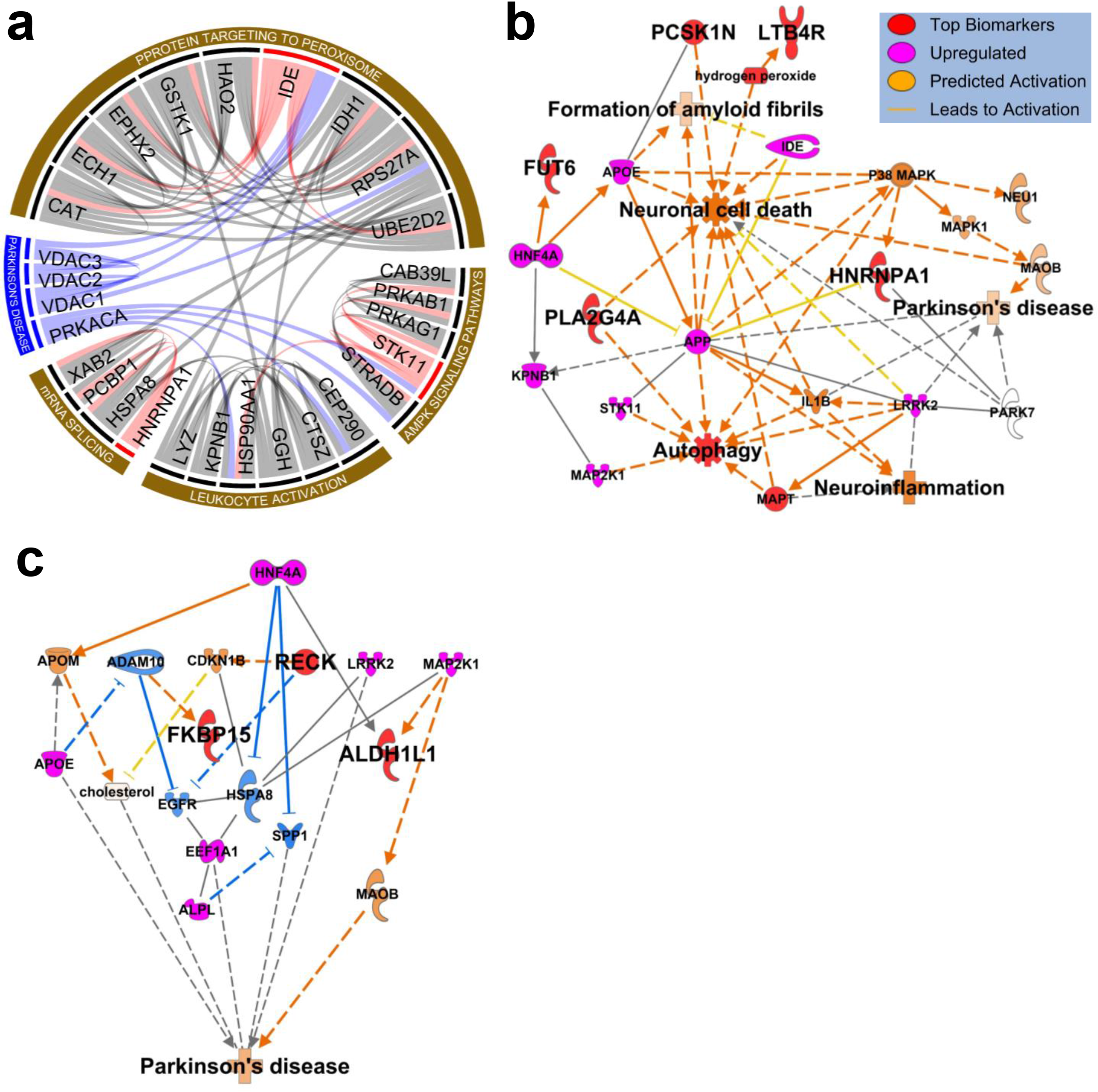
Protein and phosphoprotein biomarker network and pathway analysis. A circos plot (a) and the IPA pathway analysis of the protein and phosphoprotein disease markers (b) and risk markers (c). See **Supplementary Fig. 11** for a complete figure legend.

Additionally, **Figure 4c** depicts the pathways on how risk markers, ALDH1L1, pRECK, and pFKBP15 proteins, could trigger PD. The upregulated MAP2K1 causes the upregulation of ALDH1L1 and MAOB, which further activates PD. Selegiline, an inhibitor of MAOB protein, has been approved by the FDA to treat PD. ALDH1L1 has been developed as one of the astrocyte markers and is homogenously expressed throughout the brain^75^. From our study, this novel marker by itself appears to be sufficient enough to act as the risk biomarker for PD. The upregulation of APOE inhibits the expression of ADAM10; consequently, pFKBP15 is overexpressed, and EGFR expression is downregulated.

Meanwhile, pRECK overexpression also inhibits EGFR expression. EGFR downregulation has been shown in postmortem brains of patients^76^. HNF4A can inhibit SPP1 and activate APOM, which leads to HDL-cholesterol activation. High-plasma HDL is associated with increased PD risk and duration^77, 78^. The overexpressed ALPL also inhibits SPP1, a glycosylated phosphoprotein expressed in neurons, and appears to play the role of a double-edged sword in neurodegenerative diseases^79^. SPP1 may be toxic to neurons, lead to cell death in some cases, and have potent neuroprotective effects in others^80^.

### Top disease and risk biomarkers were selected and evaluated using ten-fold cross-validation

The discovery experiment, which included samples from 21 healthy individuals (control), 13 healthy individuals with G2019S mutation in the *LRRK2* gene (non-manifesting carrier/NMC), 28 PD individuals without G2019S mutation (idiopathic PD/iPD), and 20 PD individuals with G2019S mutation (LRRK2 PD), were randomly divided into two groups: 70% training set and 30% test set for biomarker selection and predictive ability estimation (**Figure 1**). The median normalization was performed on the training set so that all abundances in the four groups had the same median. After passing thresholds and robust normalizations, we obtained and quantified a total of 2,127 qualified unique proteins and 1,153 qualified unique phosphoproteins.

From these curated training data, we generated six volcano plots for comparisons between NMC, iPD, and LRRK2 PD groups against the control samples with cut-off values of unpaired Student’s two-tailed t-test p-value = 0.05 (-log10(0.05)=1.30) and log base 2 fold-change = 0.5, which equals to ∼1.414 fold-change (**Supplementary Fig. 9a and 9b**; see **Supplementary Data 12** for the proteome results, **Supplementary Data 13** for the phosphoproteome results, and **Supplementary Data 14** for overlapping proteins and phosphoproteins). Here, the volcano plots were created to facilitate the top feature selections using machine learning rather than finding significant features; therefore, a multiple hypothesis corrections were not used as the cutoff. The upregulated proteins and phosphoproteins were overlapped in Venn diagrams. As mentioned previously, we investigated potential biomarkers in two different groups, identified as disease markers and risk markers. We denoted disease markers as upregulated in PD regardless of the *LRRK2*-G2019S mutation (both iPD and LRRK2 PD groups). Risk markers were labeled as upregulated in both NMC and iPD groups (NMC and iPD groups). The upregulation of the disease biomarkers could indicate that a patient currently has PD. On the other hand, the upregulation of the risk biomarkers could mean a higher chance of developing PD in the future, especially for those NMC individuals. A single protein biomarker might be involved in several already known diseases. To offer a better diagnostic value, we proposed to quantify a set of several biomarkers rather than a single diagnostic protein.

We first performed feature selection to select the top disease and risk biomarkers. Instead of using a simple one-shot feature selection technique that usually yields a sub-optimal solution, we used a two-step feature selection process that generates better performance: backward feature elimination followed by exhaustive feature selection (See **Supplementary Table 1** for the feature selection inputs and the intermediate results after backward feature elimination and before exhaustive feature selection, see **Supplementary Data 15** for more details of feature selection inputs)^81^. We deployed backward feature elimination which removes, one feature at a time, those features that do not have a significant effect on the dependent variable or prediction of output. Then, we deployed exhaustive feature selection to find the best performing feature subset by searching across all possible feature combinations (a brute-force method), until the desired number of features is left. Specifically, this number is determined by observing the increase in performance (accuracy) with the increase in the number of final selected features (in which it is diminishing return). By utilizing this two-layer method, we could identify the top 6 disease biomarkers and 3 risk biomarkers. Disease biomarkers are upregulated in iPD and LRRK2 PD compared to HC and NMC. Meanwhile, risk biomarkers are upregulated in NMC and iPD compared to HC and LRRK2 PD. The final selected markers are shown in **Figure 5a and 5b** and listed in **Figure 5c**. The violin plots of the selected disease and risk biomarkers are shown in **Figures 5d and 5e**, respectively (see **Supplementary Fig. 10** for additional top biomarkers). Henceforth, we would label those biomarkers listed in **Figure 5c** as top biomarkers; meanwhile, we would designate those biomarkers listed in **Supplementary Table 1** but not listed in **Figure 5c** as potential biomarkers. We optimized our hyperparameters and trained our model using the random forest classifier with 10-fold cross validation by utilizing the top 6 disease biomarkers and the top 3 risk biomarkers as features in two separate analyses - one for the disease biomarkers and the other for the risk biomarkers. Lastly, we trained our model by utilizing the ten-fold cross-validation.

**Figure 5.**
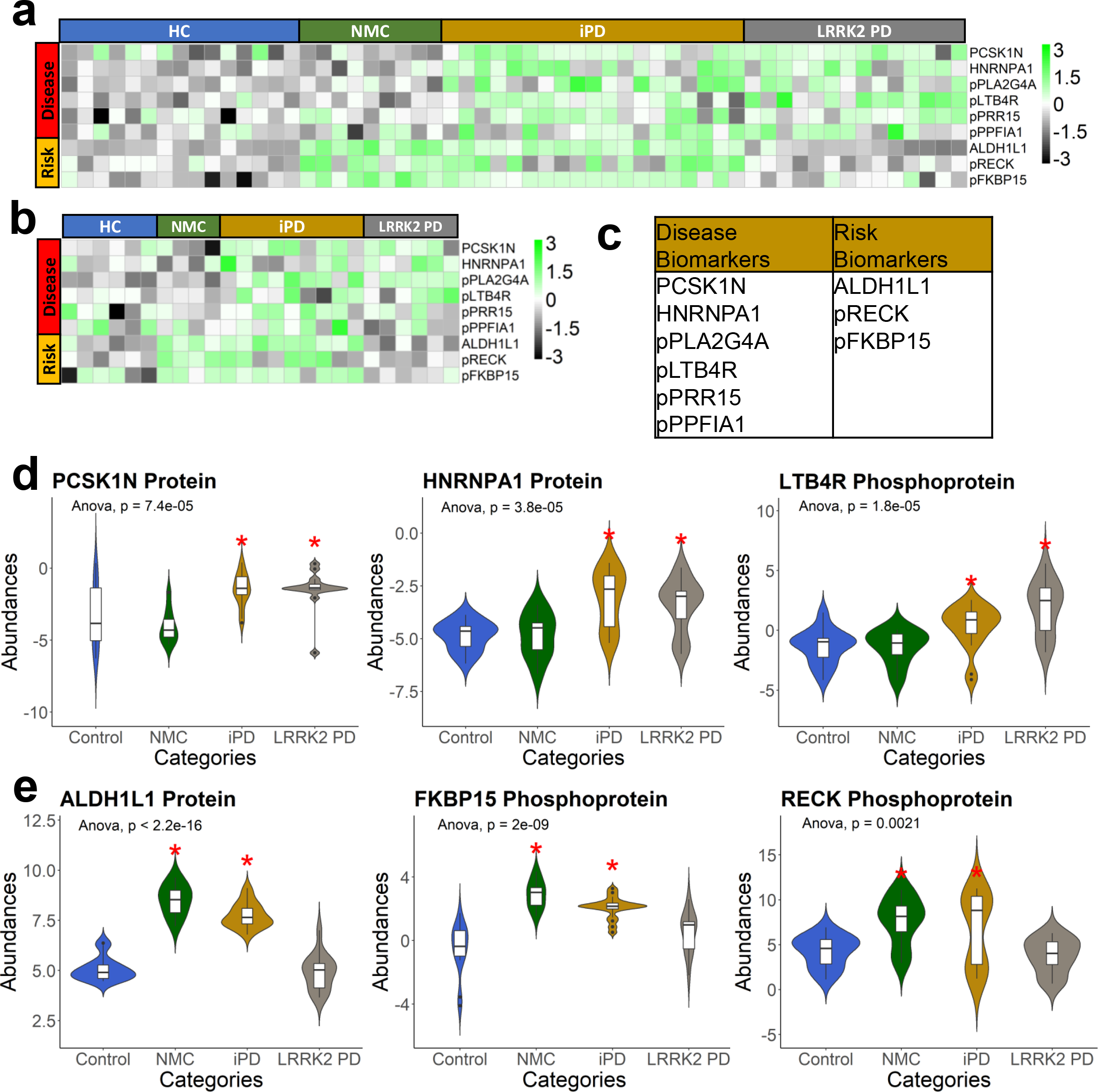
The selected top disease and risk biomarkers acquired from the training set. a) The training set’s heatmap of top potential protein and phosphoprotein biomarkers. b) The heatmap of top potential protein and phosphoprotein biomarkers on the test set. c) The table summary of the top disease and risk biomarkers. Violin plots of the statistically upregulated proteins and phosphoproteins from the training set in (d) PD regardless of the *LRRK2*-G2019S mutation (disease markers) and (e) in both NMC and iPD groups (risk markers) (see **Supplementary Fig. 10** for additional biomarkers). The unpaired two-samples Wilcoxon test p-values and the one-way ANOVA p-value were included on each of the violin plots.

### Disease biomarkers were substantiated using classification models, PRM-MS targeted mass spectrometry, and Western blot experiments

After the careful feature selection and hyperparameters as described above, we tested our constructed model using accuracy scores, confusion matrixes, and receiver operating characteristic (ROC) curves, as depicted in **Figure 6**. A confusion matrix evaluates one classifier with a fixed threshold, while the ROC evaluates that classifier over all possible thresholds. The area under the ROC curve (AUC) provides the performance measurement across the classification threshold. A higher true positive percentage and a lower false-positive percentage will produce better AUC results. Normally, in the medical field, an AUC of 70-80% is considered acceptable, 80-90% is considered good, and 90-100% is considered excellent^82^. For example, the AUC for the top six disease biomarkers is 94.3%, with 87.60% confusion matrix accuracy (**Figure 6a and 6c**). This panel would result in a 94.3% likelihood that the doctor will correctly distinguish a PD patient from a healthy patient based on finding the six biomarkers at an elevated level in the urinary EVs. Furthermore, we found that using all three protein and phosphoprotein risk biomarkers resulted in an AUC of 99.80% with 96.56% accuracy for the confusion matrix (**Figure 6b and 6d**). Certainly, these findings need to be verified with a much more expanded validation cohort.

**Figure 6.**
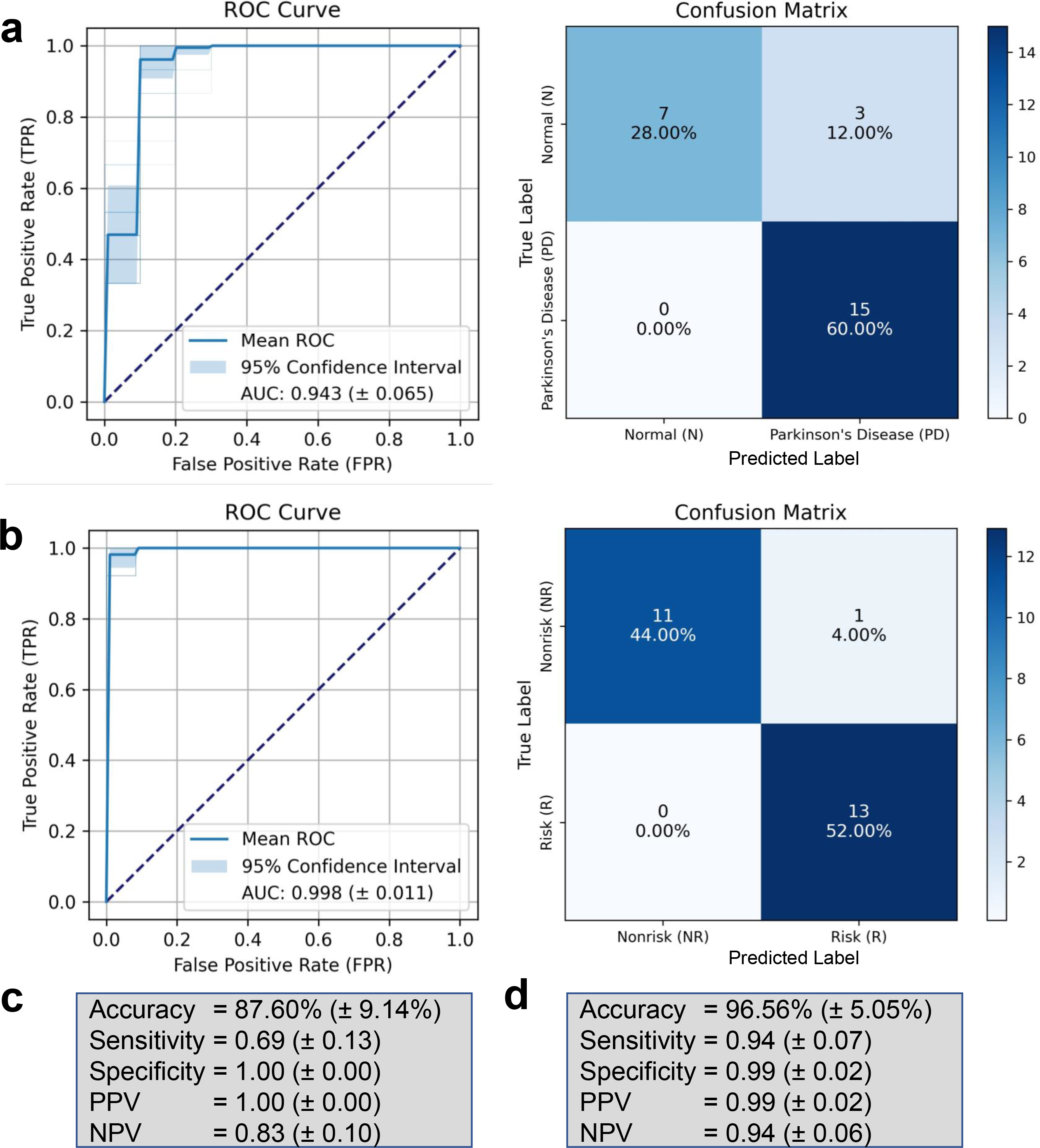
Unbiased estimation of predictive ability of urinary proteomes and phosphoproteomes on the test set. Receiver operating characteristic (ROC) curves and the confusion matrixes for the Random Forest Classifier model to classify (a) Parkinson’s Disease (PD) vs. Normal (N) individuals and (b) Risk (R) vs. Nonrisk (NR) individuals. The dotted diagonal line indicates random performance, and the light blue area represents the 95% confidence interval. The accuracy, sensitivity, specificity, positive predictive value (PPV), and negative predictive value (NPV) scores with their 95% confidence intervals are shown for both (c) the PD vs. N and (d) the R vs. NR.

Parallel Reaction Monitoring-Mass Spectrometry (PRM-MS) and Western blot are both commonly used for the initial validation of potential biomarkers. We selected several of our top disease biomarkers and urinary exosome markers for further validation. Quantitative assays based on PRM-MS for the disease markers were performed with a new set of urine EV samples from 23 patients with PD and 13 healthy individuals (see **Supplementary Data 7** for the cohort demographics and clinical characteristics). The risk markers were also validated using PRM-MS from the same urine EV samples (22 risk patients and 14 non-risk individuals). All of the samples used in the validation experiments came from a new cohort of patients. In this PRM-MS experiment, we targeted several top EV markers, proteins involved in PD pathways, proteins known to be PD biomarkers, several potential disease and risk biomarkers (those biomarkers which were not chosen as top biomarkers by a feature selection), and finally the top disease and risk biomarkers (see **Supplementary Table 2** for the target list, see **Supplementary Data 16** for the inclusion list). Due to limited volumes of the new urine samples, we could only target protein biomarkers.

One out of two targeted top disease biomarkers, HNRNPA1, was observed to be significantly upregulated in patients with PD compared to healthy individuals (**Figure 7a**, see **Supplementary Data 16** for the table format). HNF4A, whose mRNAs were found to be upregulated in the blood of 51 PD patients vs. 45 controls using quantitative PCR assays, was significantly upregulated in PD (P < 0.01). Meanwhile, APP, whose mRNAs have been shown as blood biomarkers of PD, was also significantly upregulated in PD (P < 0.05, **Supplementary Fig. 11**)^83^. Interestingly, the upregulation of CD9, CD63, and CD81 agreed with the previous finding that these three EV markers’ median fluorescence intensity (MFI) on the surface of plasma-derived EVs was significantly higher in PD compared to HC (P < 0.05)^84^. FN1, one of the proteins normally found to be co-purified with EVs, was also upregulated in PD patients (P < 0.01). Furthermore, the top protein risk biomarker, ALDH1L1, was successfully validated along with two potential risk biomarkers, ALPL and B4GALT3 (P < 0.05).

**Figure 7.**
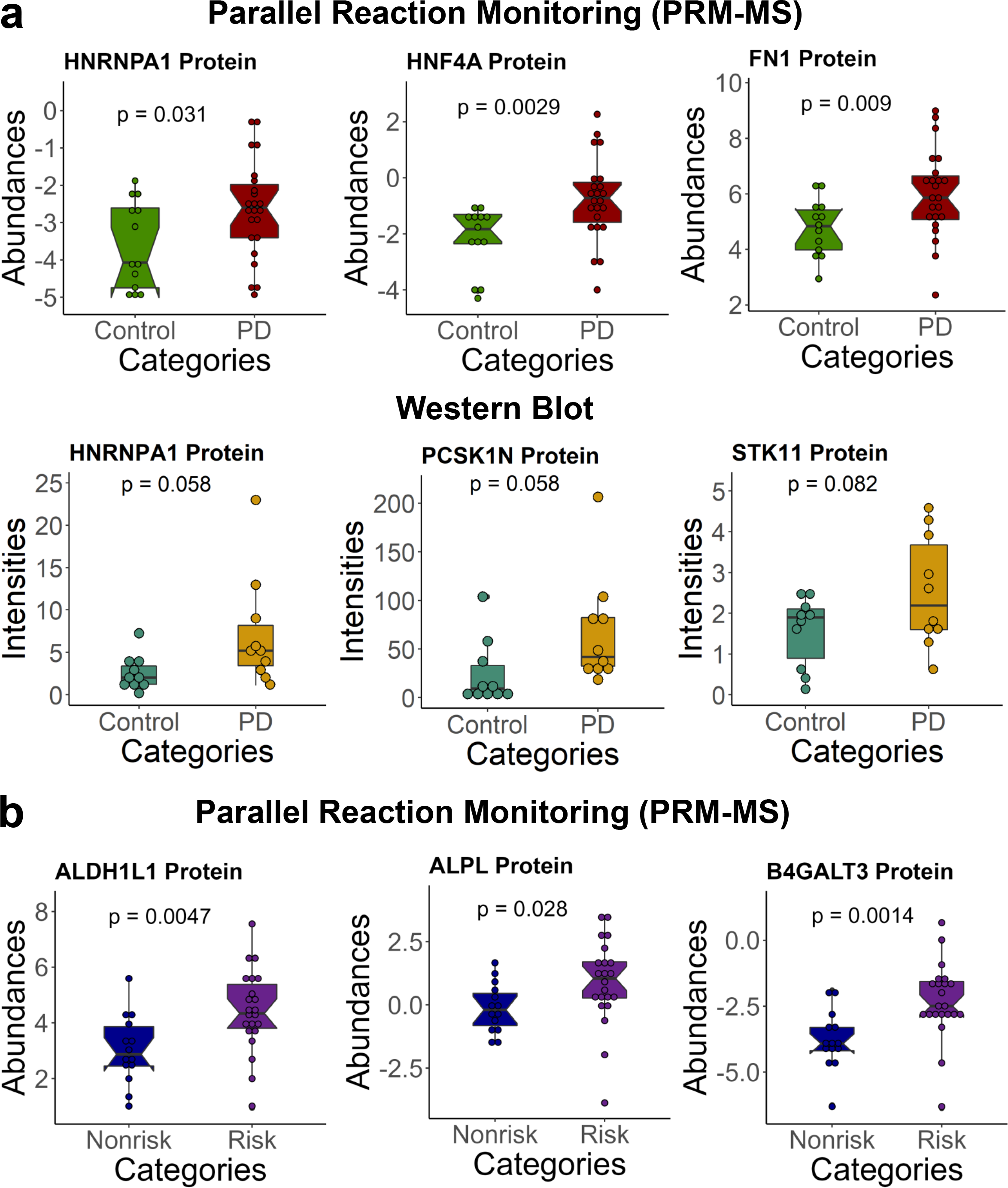
Targeted quantitation of disease and risk biomarkers. a) A top disease biomarker, HNRNPA1, was validated in 23 patients with PD and 13 healthy individuals, using PRM-MS (P < 0.05). HNF4A and FN1 were also significantly upregulated in PD (P < 0.01). Two top disease biomarkers, HNRNPA1 and PCSK1N, and a potential disease biomarker, STK11, were validated in 10 patients with PD and 10 healthy individuals using Western blot (P < 0.1). b) A top risk biomarker, ALDH1L1, was validated in 22 risk and 14 nonrisk individuals using PRM-MS (P < 0.05). Two potential risk biomarkers, ALPL and B4GALT3, were significantly upregulated (P < 0.05). The Student’s two-tailed t-test calculated all P values.

We further performed immunoassay with this new cohort of urine EV samples from 10 patients with PD and 10 healthy individuals. Three disease markers, HNRNPA1, PCSK1N, and STK11, were noticeably upregulated in patients with PD compared to healthy individuals (**Figure 7a,** see **Supplementary Fig. 12** for the original Western blot images, and see **Supplementary Data 16** for the table format).

### LRRK2 and its Rab Substrates Signaling are altered but not significant PD biomarkers

Lastly, we would like to discuss whether some Rab phosphoproteins, which are known to be direct substrates of LRRK2 and identified in EVs, would be altered across different groups in the Columbia LRRK2 cohort. Therefore, we also investigated the direct LRRK2 activation in these urine EV samples in addition to new biomarker discovery. LRRK2 is known to phosphorylate a subgroup of Rab proteins, and *LRRK2*-G2019S mutation has been previously shown to increase the phosphorylation of its Rab substrates^16^. Rab proteins are master regulators of membrane trafficking, orchestrating vesicle formation and vesicle movement along actin and tubulin networks, as well as membrane docking and fusion - all critical aspects of autophagy and lysosome biology^16^. First, we performed Western blot analyses of all 82 urine samples, quantifying CD9 (common exosome marker), LRRK2, and pSer1292-LRRK2 signal in the EVs captured by EVtrap (**Supplementary Fig. 13a,** see **Supplementary Data 17** for table format). pSer1292, a LRRK2 autophosphorylation site, indirectly reflects LRRK2 activation^85^. We normalized and compared the signal between all samples using a recombinant autophosphorylated LRRK2 protein as an internal standard^49^. As expected, the normalized CD9 signal did not show a significant difference between the sample groups (**Supplementary Fig. 13b**), while the expression of LRRK2 in LRRK2 PD was significantly higher than in the control samples (P=0.028). Unfortunately, it was challenging to detect the pSer1292-LRRK2 signal in most samples, caused by a meager amount of this modified protein in the samples and/or a lower antibody sensitivity. Due to undetectable signals in most samples, we did not find any significant difference in the pSer1292-LRRK2 phosphorylation level itself (**Supplementary Fig. 13b**). We also compared the Western blot quantitative result with the mass spectrometry data (**Supplementary Fig. 13c**). While not all of the differences observed in these Western Blot and mass spectrometry experiments showed statistically significant changes, there was an apparent trend of higher LRRK2 signal in the G2019S groups (NMC and LRRK2 PD) in both the Western blot and mass spectrometry data. Interestingly, the LRRK2 overall phosphorylation level (sites other than Ser1292) is lower in NMC and significantly lower in LRRK2 PD than in the control group. Indeed, the low level of phosphorylated LRRK2 in EVs might explain why it was challenging to detect pSer1292-LRRK2 signals in the Western Blot.

From the urine EV LC-MS analyses, we identified 34 Rab GTPases, 12 of which are known LRRK2 substrates, and eight phosphorylated Rab GTPases (**Supplementary Table 3**). After in-depth statistical normalization and qualification, we quantified 15 Rab GTPases (10 LRRK2 substrates) and four phosphorylated Rab GTPases. We observed that Rab2A (P<0.003) and Rab10 (P=0.037) were significantly upregulated in LRRK2 PD compared to the control samples (**Supplementary Fig. 14a**). Rab2A’s involvement in retrograde trafficking and particle recycling from Golgi back to the endoplasmic reticulum (ER) shows the role of this protein in the organellar homeostasis pathway to prevent misfolded proteins from entering the Golgi apparatus^6^. Thus, the upregulation of Rab2A in LRRK2 PD, which may promote retrograde trafficking machinery, may be the α-syn aggregation stress response.

Rab10 is a well-known substrate of LRRK2, and *in vitro* assays suggested that PD-related neurodegeneration may start by *LRRK2*-G2019S increasing phosphorylation of Rab10^86^. It is also known that Rab10 is involved in LRRK2 and other Rabs relocalization^66^. Therefore, it is not surprising that Rab10 was present at higher levels in the LRRK2 PD group EVs (**Supplementary Fig. 14a**). Interestingly, Rab17 protein was qualified to be one of our progression markers, although currently, the role of Rab17 in PD progression is not fully understood. At the phosphoprotein level, only Rab12 (P<0.005) was significantly upregulated in LRRK2 PD (**Supplementary Fig. 14a**). Rab12 is a LRRK2 endogenous substrate that plays a role in endosomal lysosome sorting, degradation, and autophagy^66^.

We also investigated the correlation of the identified Rab GTPase expression levels with age, gender, disease duration, and MoCA scores with the new biomarkers. As seen in **Supplementary Figure 14b**, the expression of Rab1A protein in females was significantly higher than in males with P<1e-12. In contrast, Rab1B (P<0.0005), Rab3D (P<0.005), and Rab7A (P<0.05) were expressed at higher levels in males than females. We also observed that the expression level of Rab2A protein in female iPD individuals was positively correlated with age (R^2^=0.68, P<0.001) (**Supplementary Fig. 14c**). Meanwhile, the expression of Rab17 protein in female iPD individuals was positively correlated with MoCA (R^2^=0.63, P<0.005) (**Supplementary Fig. 14d**). As noted before, these correlations need to be reproduced and evaluated further to better understand their significance.

## Discussion

Mass spectrometry (MS)-based biofluid proteome analysis and quantitation have recently gained renewed interest and excitement in disease profiling efforts. The approach offers immeasurable potential for innovative biomarker discovery. However, successful translation from MS data to human disease profiling remains limited. This limitation is partly due to the complexity of biofluids, which have a very large dynamic range and are typically dominated by a few highly abundant proteins. To date, scientists have been concentrating on finding PD biomarkers in EVs of biofluids such as CSF and plasma without paying much attention to the importance of urinary EVs as a potential source of biomarkers^87^. Here, we report in-depth analyses of proteome and phosphoproteome in urinary EVs and demonstrate the viability of developing proteins and phosphoproteins as potential disease biomarkers. We present an MS-based strategy that includes isolating EV particles from human urine utilizing EVtrap, enrichment of EV phosphopeptides, in-depth LC/MS analysis, and robust bioinformatics evaluation for biomarker discovery and qualitative verification (**Figure 1**). After we showed that our EV isolation method was reproducible and successfully depleted high-abundant free urine proteins, we analyzed EV samples from patients with *LRRK2*-G2019S mutation (NMC), idiopathic-PD (iPD), and LRRK2 PD compared to healthy individuals to identify candidate disease and risk biomarkers. In total, we identified and quantified 4,480 unique proteins from 46,240 peptide groups and 2,682 unique phosphoproteins from 10,620 phosphopeptide groups (**Figure 2a**). Then, the proteins and phosphoproteins identified were normalized and analyzed to be statistically useful for further downstream analyses (**Figure 2b and 2c**).

In total, we discovered two panels of high-confidence biomarkers, which were substantiated using ROC, machine learning, and in-depth network analysis. The disease biomarkers, PCSK1N, HNRNPA1, pPLA2G4A, pLTB4R, pPRR15, and pPPFIA1, could be employed for PD detection in a non-invasive way using a simple urine collection (**Figure 5**). Our machine learning technique generated an AUC of 94.3%, a confusion matrix accuracy of 87.60%, and a specificity of 1 for the top six disease biomarkers (**Figure 6**). Here, we successfully categorized every individual with PD in the test set as PD patients. Moreover, using all three protein and phosphoprotein risk biomarkers, ALDH1L1, pRECK, and pFKBP15, resulted in an AUC of 99.80%, a confusion matrix accuracy of 96.56%, and a specificity of 0.99. Again, we correctly classified all individuals with risk of developing PD in the respective category. The risk markers can be further investigated for early-onset PD detection. Several disease biomarkers have also been validated using targeted approaches - PRM and Western blot (**Figures 7a** and **7b**). Together, the extensive data on these potential biomarkers might serve as a future of PD detection in a non-invasive and more cost-effective manner and as a resource to the research community for further studies. In other words, this platform represents a foundational resource for the emerging field of accurate and reproducible proteomic biomarker discovery.

We also showed that some proteins and phosphoproteins sustained positive or negative correlations with gender, age, disease duration, MoCA, and UPDRS-III. Most interestingly, **Figure 3** displayed PEBP4, NEDD4L, and KLK6 with higher than 0.7 Pearson correlation scores, indicating a strong positive correlation with UPDRS-III. However, we need to emphasize that these correlations with these clinical parameters do not automatically mean causation. Some relevant correlations between the proteins and phosphoproteins and the clinical parameters should be studied further to provide more information. Furthermore, the emerging system/network analysis has revolutionized novel mechanism discovery and promising drug targets. Our literature-based network analysis of the gene expression involving these potential biomarkers has revealed the connections between our biomarkers and critical pathways that could lead to PD development. Here, we showed that the four top disease markers, PCSK1N, HNRNPA1, pPLA2G4A, and pLTB4R, are indeed involved in PD pathways such as neuronal cell death, neuroinflammation, autophagy, and formation of amyloid fibrils (**Figure 4b**).

We also directed significant attention to LRRK2 kinase and its Rab substrate proteins in urinary EVs. This project involved two groups of patients with *LRRK2*-G2019S mutation, a feature present in some PD patients. However, it is known that the mutated *LRRK2* does not necessarily lead to PD onset, and many individuals live with this mutation without developing Parkinson’s disease. This study found a minor increase in LRRK2 protein amount and its overall phosphorylation level in PD patients’ urine EVs (**Supplementary Fig. 13**). Similarly, a few select Rab proteins showed an increased EV signal in total protein amount and phosphorylation level in PD cases (Rab2A, Rab10, Rab12; **Supplementary Fig. 14**). However, none were selected as the optimal potential biomarkers for PD diagnosis. This finding further underscores the reality that Parkinson’s disease is highly complicated, with multiple signaling pathways involved in its pathology. While LRRK2 kinase is known to be involved in PD progression, detecting LRRK2 and its direct substrates in urinary EVs may not provide sufficient differentiation between cases. As carried out in this study, a more global analysis, which may or may not be directly influenced by LRRK2 activity, is needed to determine the most statistically significant biomarkers. We advocate that such a comprehensive analysis with highly stringent bioinformatics data validation gives us the best opportunity to discover the most optimal differentiating markers.

In summary, we have developed several comprehensive biomarker panels of proteins and phosphoproteins in urinary extracellular vesicles as biosignatures for Parkinson’s disease diagnosis. The study highlights our ability to isolate and identify thousands of unique proteins and phosphoproteins from relatively small volumes of urine samples by utilizing the EVtrap EV enrichment approach. These findings further validate the underlying principle that this strategy could be valuable for exploring existing resources in a wide range of diseases. Finally, we expect our immediate results, followed by extensive evaluation and validation of the new markers in the clinical setting, could improve these patients’ medical outcomes and quality of life.

## Methods

### Ethics / Patient Consent

The study was approved by Columbia University Irving Medical Center institutional review board (IRB) no. AAAP9604, and all participants signed an informed consent. The study design and conduct complied with all relevant regulations regarding the use of human study participants and was conducted in accordance with the criteria set by the Declaration of Helsinki.

### Sample Collection

All 82 urine samples used in the discovery LC-MS study and 56 urine samples used in the validation experiments were collected at Columbia University Irving Medical Center (CUIMC) and sent to our lab blindly. The samples were collected from March 2016 to April 2017 under a Michael J. Fox Foundation (MJFF)-funded LRRK2 biomarker project^32^. Each sample has been uniquely curated for *LRRK2* genotype and PD activity effects. For the initial comprehensive discovery experiments, the urine samples were collected from 21 healthy individuals (Control), 13 healthy individuals with G2019S mutation (non-Manifesting Carrier/NMC), 28 PD individuals without G2019S mutation (idiopathic-PD/iPD), and 20 PD individuals with G2019S mutation (LRRK2 PD). The 56 urine samples used for the validation experiments were classified as 33 patients with PD and 23 healthy individuals without genetic differentiation. All 138 samples were processed separately by implementing the statistical principles in experimental designs, including replication, randomization, and blocking when applicable^88^.

### EV Isolation by EVtrap

For each urine sample, approximately 10-15 mL was utilized for EV enrichment by EVtrap. Before the EVtrap capture, the urine volume was normalized based on the creatinine levels, a normalizer we found to be optimal for EV studies. The EVtrap beads were provided by Tymora Analytical and were utilized as described previously^37^. In brief, the frozen urine samples were thawed in a 37°C water bath. The samples were then centrifuged at 2,500 x g for 10 min to remove cell debris and large apoptotic bodies and diluted with EVtrap loading buffer in a 1:10 v/v ratio. The magnetic EVtrap beads were added directly to the diluted at a ratio of 20 uL EVtrap beads per 1 mL urine. The mixture was incubated for 1 hour by end-over-end rotation, and the supernatant was removed using a magnetic separator rack, the beads were washed with PBS, and the EVs were eluted by a 10 min incubation with 100 mM triethylamine (TEA, Millipore-Sigma). The eluted samples were dried entirely in a vacuum centrifuge. For Western blot analysis, the dried EV samples were lysed directly in LDS buffer (lower volumes of urine (∼0.5-2mL) were used for Western blot experiments).

### EV Isolation by Differential Ultracentrifugation (UC)

The EV isolation by UC was performed to compare with the EVtrap method to demonstrate the superior efficiency of isolating urinary EVs by EVtrap in the beginning of this study. Urine samples (∼1-2mL) were centrifuged at 10,000 × g at 4°C for 1 h. Supernatants were further centrifuged at an ultra-high speed of 100,000 × g (Optima MAX-XP Ultracentrifuge, Beckman Coulter) at 4°C for 2 hrs. Pellets were washed with 1x PBS and centrifuged at 100,000 × g for 2 hrs again. Collected pellets were lysed directly in LDS buffer for Western blot analysis.

### Western Blot Experiments

A small percentage (approximately 0.5 mL urine sample equivalent for CD9, 1 mL for LRRK2, and 2 mL for pSer1292-LRRK2) of each purified EV sample was incubated for 10 min at 95°C in LDS sample buffer. The equivalent volume of each sample aliquot was loaded and separated on an SDS-PAGE gel (NuPAGE 4-12% Bis-Tris Gel, Thermo Fisher Scientific). Afterward, the proteins were transferred onto a low-fluorescence PVDF membrane (Millipore-Sigma), and the membrane was blocked with 1% BSA in TBST for 1 hr. The membranes were then incubated with rabbit anti-CD9 (clone D3H4P; Cell Signaling Technology) at 1:5,000 ratio, or anti-LRRK2 (clone MJFF2 (c41-2); Abcam) at 1;1,000 ratio, or anti-pSer1292-LRRK2 (clone MJFR-19-7-8; Abcam) at 1:500 ratio overnight in 1% BSA in TBST (3% BSA in TBST was used for anti-pSer1292-LRRK2). For the secondary antibody visualization, Goat anti-Rabbit Alexa-Fluor 800 nm (Thermo Fisher Scientific) was used to bind the primary antibodies and incubated for 1 hr in 1% BSA in TBST. Lastly, the membrane was scanned by Odyssey near-infrared scanner (Licor) for signal detection and quantitation. A total of 8 blots were used for each protein target detection. We loaded internal standards at an identical concentration in each blot to normalize the signal between the samples and the blots. For CD9 relative quantitation, we extracted EVs from a mixture of several unrelated samples as an internal control added as a separate lane into each gel to enable gel-to-gel relative quantitation of the signal. For the relative quantitation of LRRK2, we used the same amount of the recombinant LRRK2 protein purchased from Thermo Fisher as an internal control for gel-to-gel relative quantitation of signal. Finally, for pSer1292-LRRK2 relative quantitation, we carried out in vitro autophosphorylation assay of the purchased recombinant LRRK2 protein, as described previously, and loaded the phosphorylated protein as an internal control for all phospho-LRRK2 Western Blot detection experiments.

For the validation experiments, the membranes were incubated with the following primary antibodies: rabbit anti-CD9 (clone D3H4P; Cell Signaling Technology) at 1:5,000 ratio, rabbit anti-STK11 (clone D60C5; Cell Signaling Technology) at 1:1,000 ratio, or mouse anti-PCSK1N (clone NP_037403.1; Millipore-Sigma) at 1:1,000 ratio, or rabbit anti-HNRNPA1 (clone D21H11; Cell Signaling Technology) at 1:1,000 ratio. For the secondary antibody visualization, Goat anti-Rabbit or Goat anti-Mouse Alexa-Fluor 800 nm (Thermo Fisher Scientific) were used to bind the primary antibodies. An equal amount of pooled urine EVs was loaded in lane 1 of each gel to normalize the signal between two blots. The signal for each sample was then normalized to CD9.

### LC-MS Sample Preparation

Phase-transfer surfactant (PTS) aided procedure was used to lyse the dried EVs and extract proteins^89^. First, EVs were resuspended in the lysis solution containing 12 mM sodium deoxycholate (SDC; Sigma-Aldrich, cat. no. D6750), 12 mM sodium lauroyl sarcosinate (sarkosyl; Sigma-Aldrich, cat. no. L9150), 10 mM TCEP-HCl (Sigma-Aldrich, cat. no. C4706), 40 mM CAA (Sigma-Aldrich, cat. no. C0267), and phosphatase inhibitor cocktail (Millipore-Sigma, cat. no. P2850) in 50 mM Tris·HCl, pH 8.5 (Tris base; Fisher BioReagents, cat. no. BP152-1 and HCl; Fisher Chemical, cat. no. A144SI-212) by incubating 10 min at 95°C. During this step, the proteins were also denatured, reduced, and alkylated. The samples were diluted fivefold with 50 mM triethylammonium bicarbonate and digested with Lys-C (Wako Chemicals, cat. no. 129-02541) at 1:100 (wt/wt) enzyme-to-protein ratio for 3 h at 37°C. For further protein digestion, trypsin (proteomics grade, modified; Sigma-Aldrich, cat. no. T6567) was added to a final 1:50 (wt/wt) enzyme-to-protein ratio for overnight digestion at 37°C. Then, the samples were acidified with trifluoroacetic acid (TFA; Merck, cat. no. 8082600100) to a final concentration of 1% TFA. An ethyl acetate (Fisher Chemical, cat. no. E145-4) solution was added at a 1:1 ratio to the samples. The mixture was vortexed for 2 min and then centrifuged at 20,000 × g for 2 min to obtain aqueous and organic phases. The organic phase (top layer) was removed, and the aqueous phase was collected, dried down to <10% original volume in a vacuum centrifuge, and desalted using TopTip C18 tips (Glygen Corporation, cat. no. TT2C18) according to the manufacturer’s instructions. After desalting, the peptide concentrations were determined by the Pierce Quantitative Peptide Colorimetry assay (ThermoFisher, cat. No. 23275), and the samples were further normalized. Each sample was split into 99% and 1% aliquots for phosphoproteomic and proteomic experiments, respectively. The samples were dried entirely in a vacuum centrifuge and stored at −80°C. For phosphoproteome analysis, the 99% portion of each sample was subjected to phosphopeptide enrichment using PolyMAC Phosphopeptide Enrichment kit (Tymora Analytical Operations, cat. no. 707) according to manufacturer’s instructions, and the eluted phosphopeptides dried completely in a vacuum centrifuge. The whole enriched sample was loaded onto LC-MS for phosphoproteomics analysis, while only 50% of each sample (equivalent to 0.5µg) was injected for proteomics.

### LC-MS Analysis

Both proteomic and phosphoproteomic samples were spiked with an 11-peptide indexed Retention Time internal standard mixture (Biognosys) to normalize the LC-MS signal between the samples. All samples were captured on a 2-cm Acclaim PepMap trap column (PN 164535, Thermo Fisher Scientific) and separated on a heated 50-cm Acclaim PepMap column (PN 164942, Thermo Fisher Scientific) containing C18 resin. The mobile phase buffer consisted of 0.1% formic acid (FA; Sigma-Aldrich, cat. no. F0507) in HPLC grade water (buffer A) with an eluting buffer containing 0.1% formic acid in 80% (vol/vol) acetonitrile (ACN; Fisher Scientific, cat. no. A955-4) (buffer B) run with a linear 60-min gradient of 6–30% buffer B at a flow rate of 300 nL/min. The UHPLC was coupled online with a Q-Exactive HF-X mass spectrometer (Thermo Fisher Scientific). The mass spectrometer was run in the data-dependent mode, in which a full-scan MS (from m/z 375 to 1,500 with the resolution of 60,000 at *m/z* 200) was followed by MS/MS of the 15 most intense ions (30,000 resolution at *m/z* 200; normalized collision energy - 28%; automatic gain control target (AGC) - 2E4, maximum injection time - 200 ms; 60sec exclusion].

### Parallel Reaction Monitoring (PRM)

Peptide samples were dissolved in 10.8 μL 0.05% TFA & 2% ACN and injected 10 μL into the UHPLC coupled with a Q-Exactive HF-X mass spectrometer (Thermo Fisher Scientific). The mobile phase buffer consisted of 0.1% formic acid in HPLC grade water (buffer A) with an eluting buffer containing 0.1% formic acid in 80% (vol/vol) acetonitrile (buffer B) run with a linear 60-min gradient of 5–35% buffer B at a flow rate of 300 nL/min. Each sample was analyzed under PRM with an isolation width of ±0.8 Th. In these PRM experiments, an MS2 level at 30,000 resolution relative to *m/z* 200 (AGC target 2E5, 200 ms maximum injection time) was run as triggered by a scheduled inclusion list. The inclusion list included peptides that have been manually picked and compared to PeptideAtlas^90, 91^. Higher-energy collisional dissociation was used with 28 eV normalized collision energy. PRM data were manually curated within Skyline-daily (64-bit) 22.1.9.208 (6839020bd)^92^.

### LC-MS Data Processing

The raw files were searched directly against the human Swiss-Prot database with no redundant entries, using Byonic (Protein Metrics) and Sequest search engines loaded into Proteome Discoverer 2.3 software (Thermo Fisher Scientific). MS1 precursor mass tolerance was set at 10 ppm, and MS2 fragment tolerance was set at 20 ppm. In the processing workflow, search criteria for both search engines were performed with full trypsin/P digestion, a maximum of two missed cleavages allowed on the peptides analyzed from the sequence database, a static modification of carbamidomethylation on cysteines (+57.0214 Da), and variable modifications of oxidation (+15.9949 Da) on methionine residues and acetylation (+42.011 Da) at N terminus of proteins. Phosphorylation (+79.996 Da) on serine, threonine, or tyrosine residues was included as the variable modification for phosphoproteome analysis. The false-discovery rates of proteins and peptides were set at 0.01. All protein and peptide identifications were grouped, and any redundant entries were removed. Unique peptides and unique master proteins were reported. Finally, the proteomic results were further normalized against common urine EV proteins to account for any other variations in urine concentration.

### Label-free Quantitation Analysis

The label-free quantitation node of Precursor Ions Quantifier in the consensus workflow through the Proteome Discoverer v2.3 (Thermo Fisher Scientific) was used to quantify all data. For the quantification of proteomic and phosphoproteomic data, the intensities of peptides were extracted with initial precursor mass tolerance set at 10 ppm, fragment mass tolerance at 0.02 Da, minimum peak count as 1, maximum RT shift as 5 min, PSM confidence FDR of 0.01 as strict and 0.05 as relaxed, with hypothesis test of t-test (background based), protein abundance based ratio calculation, 100 as the maximum allowed fold change, and site probability threshold of 75. The abundance levels of all peptides and proteins were normalized to the spiked-in internal iRT standard. For calculations of protein abundance, the sum of sample abundances of the connected peptide groups was added together and used for downstream analysis.

### Bioinformatics Analysis

All clinical sample data were analyzed using the Perseus software (version 1.6.5.0)^93^. The normalized intensities of proteins and phosphoproteins were extracted from Proteome Discoverer search results and log-based 2 transformed for quantifying both proteomic and phosphoproteomic data. The abundances were categorized into four different categories: Control, NMC, iPD, and LRRK2 PD. The proteins or phosphoproteins with detected abundances of more than 70% in each category were kept. It was done to keep the proteins and phosphoproteins detected in at least one category. The imputation for the missing abundances was performed by assigning small random values from the normal distribution with a downshift of 1.8 SDs and a width of 0.3 SDs. Very low abundances normally cause missing values.

All abundances for each protein or phosphoprotein were further normalized by subtracting the median from each protein or phosphoprotein abundance. Then, the unpaired unpaired Student’s two-tailed t-test was performed, and the difference in averages was calculated for the three comparisons. Various packages in R 3.5.0^94^, including but not limited to ggplot2 3.3.1^95^, ggpubr 0.3.0^96^, EnhancedVolcano 1.7.6^97^, pROC^98^, Vennerable 3.0^99^, and Circlize 0.4.9^100^, and also Cytoscape 3.8.0^101^ (an open-source software platform for visualizing complex networks) were used to visualize the data. For the volcano plots, the x-axis is the log(2) fold-change on averages, and the y-axis is the log(10) of the p-value. Volcano plots were created for each of comparison with cut-off values of permutation-based FDR = 0.05 (-log10(0.05)=1.30) and log base 2 fold-change = 0.5, which equals to ∼1.414 fold-change. The Venn diagrams were created based on the upregulated proteins or phosphoproteins in the volcano plots. The violin plots were generated by focusing on significant proteins and phosphoproteins from the overlapped area in the Venn diagrams, which represented 2 different categories: disease markers and risk markers. The unpaired two-samples Wilcoxon test p-values and the one-way ANOVA p-value were included on each of the violin plots. The unpaired two-samples Wilcoxon test p-values were shown for the comparison between gender and specific proteins. The correlations between potential biomarker expressions with gender, age, disease duration, and MoCA were created with a minimal 0.6 for R^2^ and a maximal 0.05 for p-value calculated using t-distribution with n-2 degrees of freedom as thresholds. Lastly, STRING v11.5^59^ and IPA^102^ (QIAGEN Inc., https://www.qiagenbioinformatics.com/products/ingenuitypathway-analysis) were used to analyze the protein-protein interactions and validate their respective protein roles in hallmark PD pathways.

### Division Into Training Set and Test Set, Feature Selection, and Predictive Analyses

One hundred forty-one unique subjects were divided randomly into the discovery and validation experiments. In total, 82 subjects were categorized into the main experiment, further divided into training (57 subjects) and test sets (25 subjects). Fifty-nine subjects were used for the validation experiments, consisting of parallel reaction monitoring and Western Blot experiments. To facilitate the top feature selection using the machine learning, we created volcano plots for each of comparison in the training set with cut-off values of unpaired Student’s two-tailed t-test p-value = 0.05 (-log10(0.05)=1.30) and log base 2 fold-change = 0.5, which equals to ∼1.414 fold-change (See **Supplementary Data 12 and 13** for the volcano plot results). For the discovery experiment, we first performed feature selection on the biomarker candidates listed on **Supplementary Data 14** obtained from **Supplementary Figure 9**. The goal was to discover both disease and risk biomarkers in an independent manner. Disease biomarkers are upregulated in iPD and LRRK2 PD compared to HC and NMC. Meanwhile, risk biomarkers are upregulated in NMC and iPD compared to HC and LRRK2 PD. For the feature selections, machine learning, and predictive analyses, we utilized various packages, such as python 3.8.8^103^, conda 4.13.0^104^, jupyter-notebook 6.3.0^105^, pandas 1.4.3^106^, numpy 1.20.1^107^, matplotlib 3.3.4^108^, plotly 5.6.0^109^, sklearn 1.1.1^110^, mlxtend 0.20.0^111^, and xgboost 1.6.1^112^. Instead of using a simple one-shot feature selection technique that usually yields a sub-optimal solution, we used a two-step feature selection process that generates better performance: backward feature elimination followed by exhaustive feature selection^81^. We deployed backward feature elimination which removes, one feature at a time, those features that did not have a significant effect on the dependent variable or prediction of output. Then, we deployed exhaustive feature selection, which aims at finding the best performing feature subset by searching across all possible feature combinations (a brute-force method) until the desired number of features is left. Specifically, this number was determined by observing the increase in performance (accuracy) with the increase in the number of final selected features (in which it is diminishing return). For the 48 potential disease biomarkers, we performed feature selection as follows: 48 features -> backward feature elimination -> 14 features -> exhaustive feature selection -> 6 features. Meanwhile, for the 22 potential risk biomarkers, we selected the features as follows: 22 features -> backward feature elimination -> 10 features -> exhaustive feature selection -> 3 features. At the end of the feature selection, we discovered a list of top 6 disease biomarkers and top 3 risk biomarkers.

Next, we performed a hyperparameters selection process using which included a randomized search followed by an exhaustive search on a random forest classifier with 10-fold cross validation utilizing the top 6 disease biomarkers and the top 3 risk biomarkers in two separate analyses. In particular, we searched over the following set of hyperparameters: ‘n_estimators,’ ‘max_features,’ ‘max_depth,’ ‘min_samples_split,’ ‘min_samples_leaf,’ and ‘bootstrap’ in which we validated the result by using ten-fold cross-validation.

The best set of hyperparameters for disease biomarkers is as follows: ‘bootstrap’: True, ‘max_depth’: 4, ‘max_features’: ‘auto’, ‘min_samples_leaf’: 3, ‘min_samples_s plit’: 3, ‘n_estimators’: 320. Meanwhile, the best set of hyperparameters for risk biomarkers is as follows: ‘bootstrap’: True, ‘max_depth’: None, ‘max_features’: ‘auto’, ‘min_samples_leaf’: 3, ‘min_samples_split’: 3, ‘n_estimators’: 240.

In randomized search, we searched across 200 different combinations of hyperparameters and then created the hyperparameter grid encompassing the optimal sampled hyperparameter combination from the randomized search. An exhaustive search was used to select the best performing set of hyperparameters from the generated grid. Finally, we tested our constructed model (with carefully chosen features and hyperparameters described above) 50 times and evaluated it by considering the accuracy, confusion matrix, and ROC curve.

Finally, we repeatedly trained a Random Forest Classifier with the selected features and hyperparameters obtained from the above processes 50 times. After that, we evaluated each constructed model using accuracy, confusion matrix, and ROC curve. To summarize the results over all trials, we computed each evaluation metric’s mean and 95% confidence interval.

### Data Availability

The mass spectrometry raw data files and Proteome Discoverer search results have been deposited in the MassIVE database (https://massive.ucsd.edu/ProteoSAFe/static/massive.jsp) and can be accessed via dataset identifier: MSV000085800 | PXD020475. They were made available to the reviewers.

The PRM raw data files and the Skyline file have been deposited in the Panorama Public (https://panoramaweb.org/xdfV55.url). They were made available to the reviewers.

### Code Availability

Custom Python code used for feature selections, model training, and predictive analyses and the README file was made available to the reviewers and will be made available to the public upon the manuscript publication.

## Data Availability

All data produced in the present study are available upon reasonable request to the authors.

## Acknowledgements

This project was partially supported by The Michael J. Fox Foundation for Parkinson’s Research (Grant ID 17453) and NIH grant (Grant ID 3RF1AG064250). We also thank Dr. Andrew West and Shijie Wang for their protocol suggestions for LRRK2 autophosphorylation and p-LRRK2 detection on Western Blot. Cartoon in **Figure 1** created with BioRender.com.

## Contributions

A.I. and L.L. performed the experiments and analyzed the data. M.H. carried out the bioinformatics and statistical analyses, and generated the figures, final data, and markers. M.H. and K.K. designed, optimized, and executed the machine learning. X.W. conducted the urine normalization and optimized the PRM method. M.H. and Z.L. designed and created **Figure 1** with BioRender.com. R.N.A. provided the urine samples and information about the cohort. A.I., S.P, and W.A.T. conceived the project and designed the experiments. M.H., A.I., and W.A.T. wrote the paper. All authors reviewed and approved the manuscript.

---

**Supplementary Figure 1.**
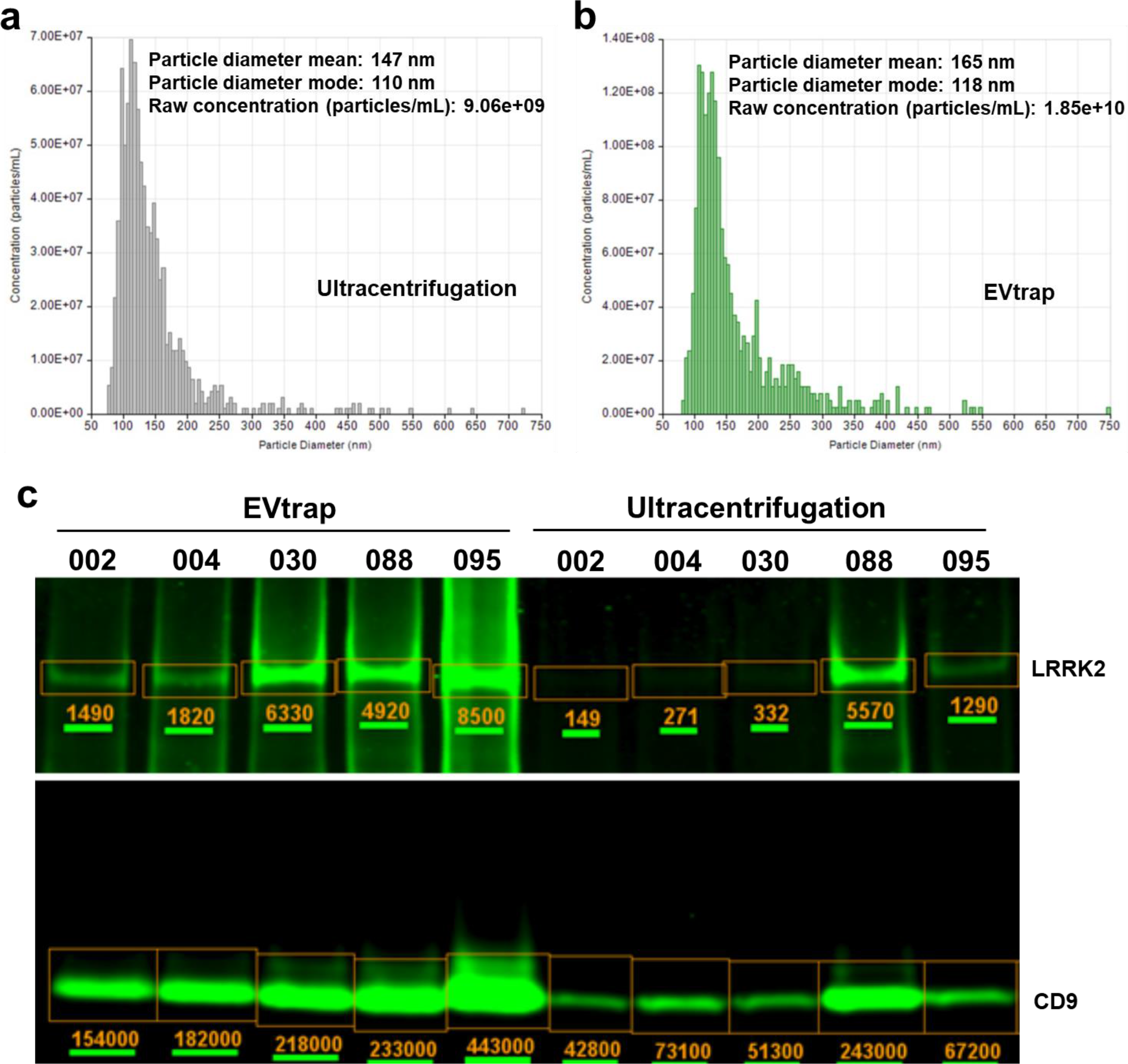
Comparison of EVtrap and ultracentrifugation for EV capture from urine. TRPS analysis of EVs captured by a) ultracentrifugation or b) EVtrap. c) Western blot detection of CD9 and LRRK2 proteins of 5 urine EV samples isolated by EVtrap or ultracentrifugation.

**Supplementary Figure 2.**
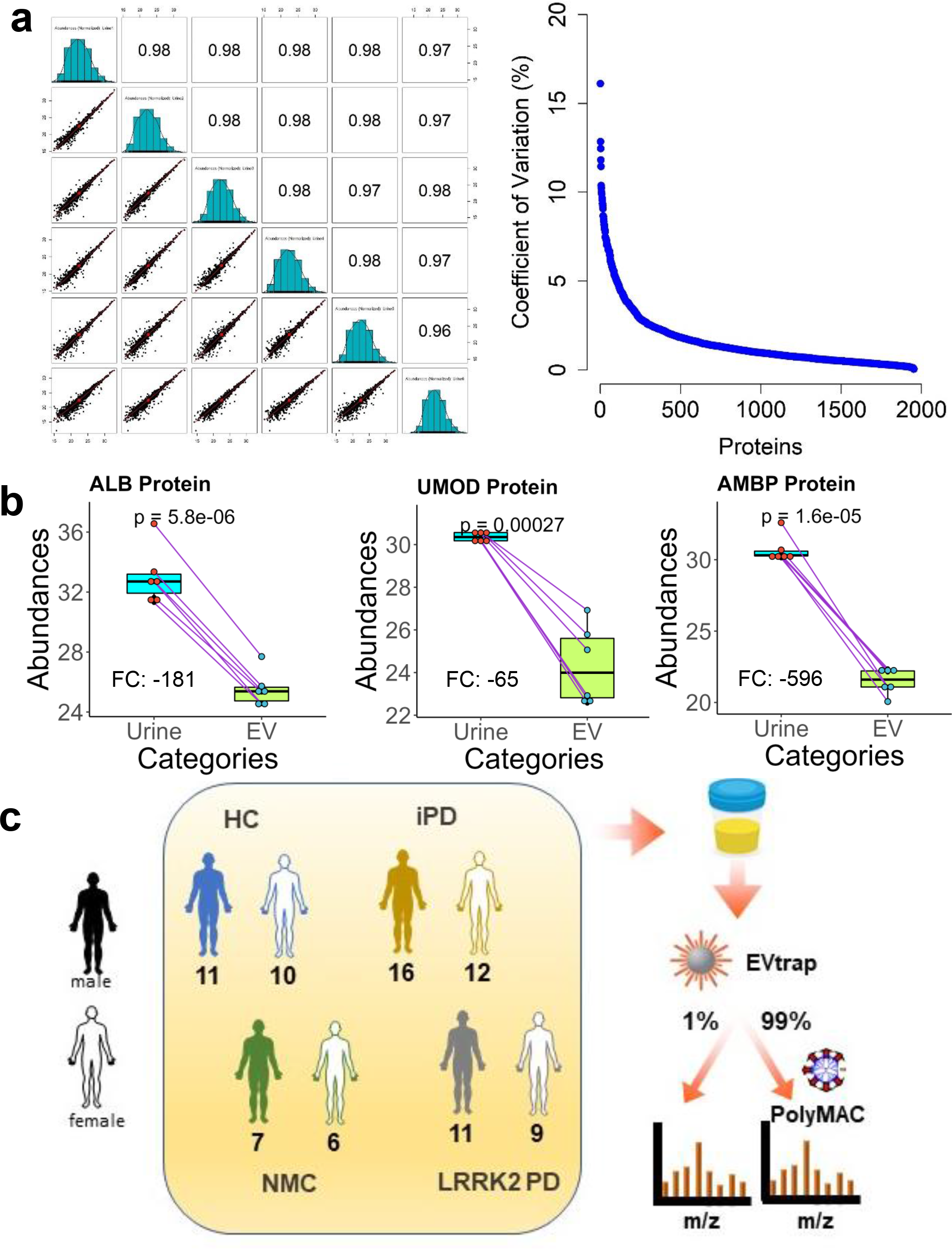
The reproducibility evaluation of our complete urine EV analysis protocol and the analytical sample preparation workflow. To evaluate the procedural reproducibility, a single urine sample was separated into six aliquots and processed with our EVtrap-LCMS protocol as six technical replicates. a) We created a multi-scatter plot accompanied by Pearson correlation coefficients and a distribution plot of proteins by a coefficient of variation (%). b) The high-abundant free urine protein depletion after urinary EVs isolation (paired Student’s two-tailed t-test p-values were shown). c) Workflow for urine samples processing.

**Supplementary Figure 3.**
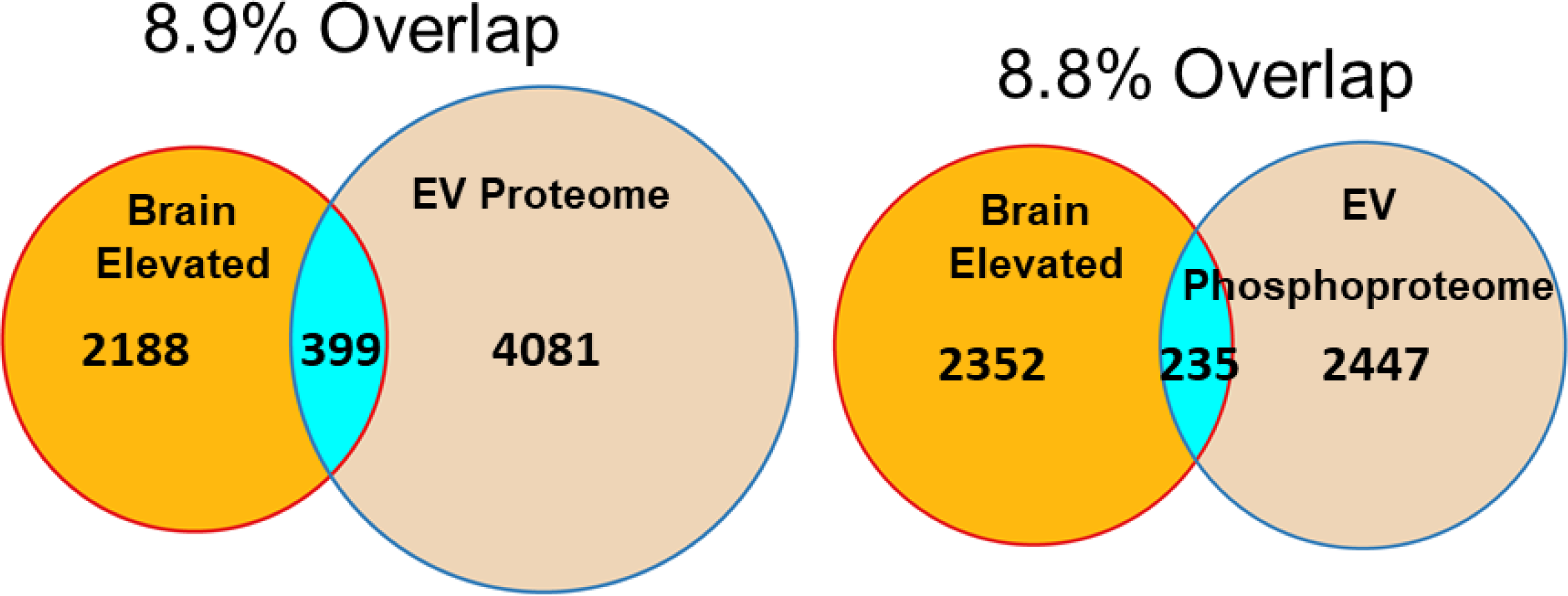
The overlap of our identified proteomic and phosphoproteomic data with available brain-elevated RNA-seq data downloaded from the Human Protein Atlas website. We used 2587 proteins classified as brain-elevated from the Human Protein Atlas for comparison with our identified EV proteins and phosphoproteins.

**Supplementary Figure 4.**
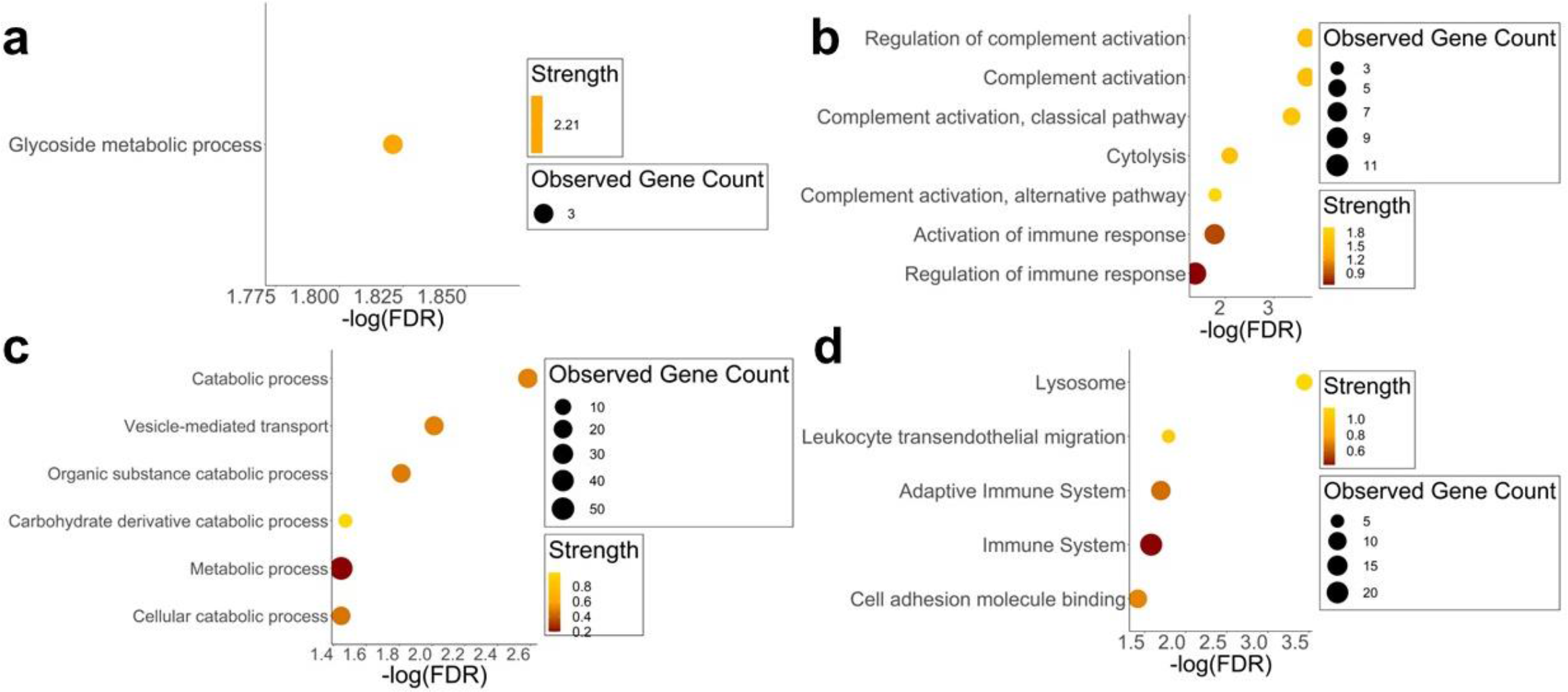
Enriched biological process gene ontology analyses of up-regulated proteins. GO analyses for a) NMC compared to control; b) iPD compared to control; c) LRRK2 PD compared to control; d) LRRK2 PD compared to iPD samples. The analyses were carried out with the STRING database. The gene ontology analyses were set with a threshold FDR of 5% after Benjamin-Hochberg correction.

**Supplementary Figure 5.**
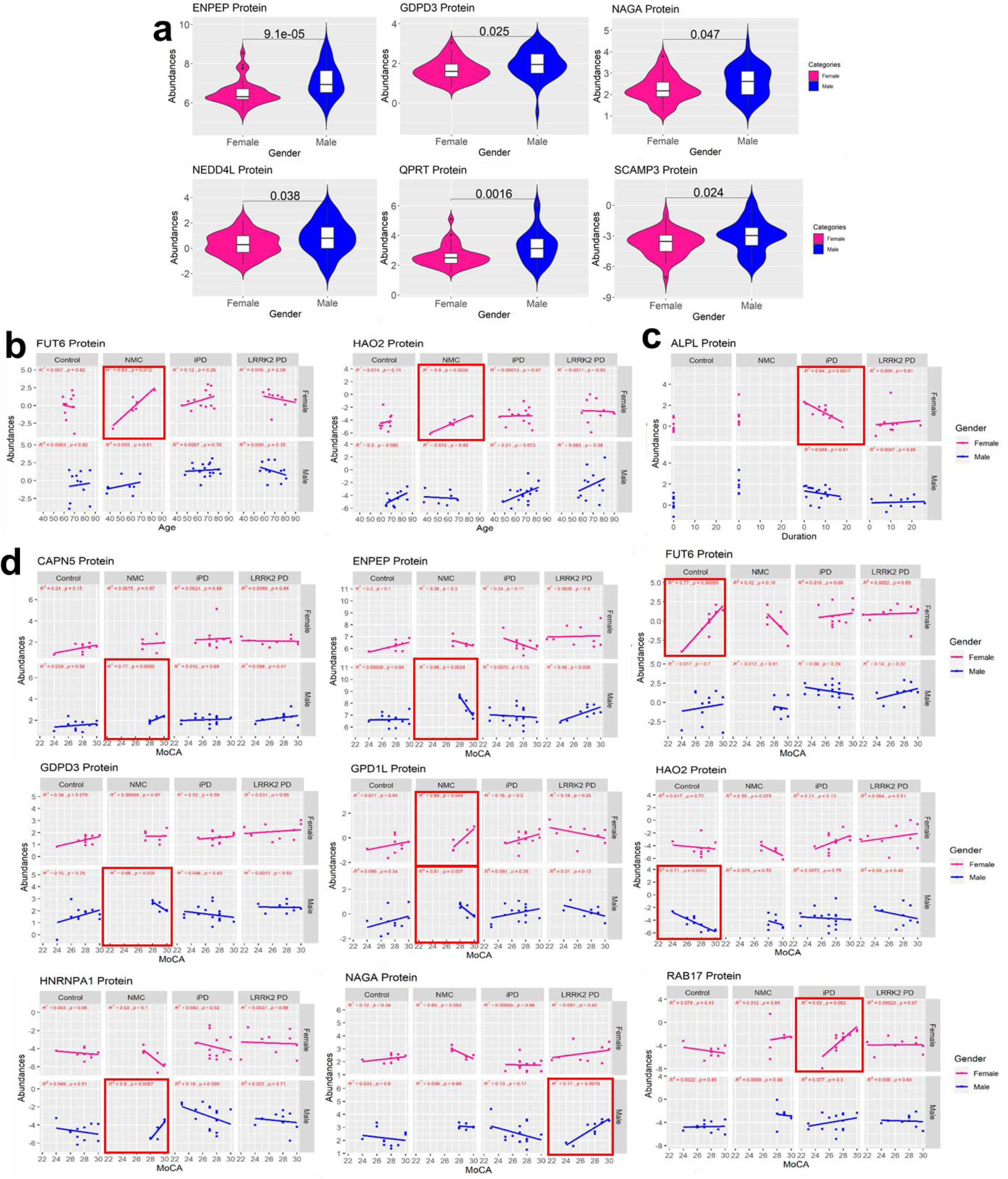
Correlation analysis for select potential protein biomarkers from the full data set (training and test sets). a) ENPEP, GDPD3, NAGA, NEDD4L, QPRT, and SCAMP3 proteins were expressed higher in males. b) The correlation analysis between FUT6 and HAO2 protein abundances and age for each group according to gender. c) The correlation analysis between ALPL protein abundances and disease duration for each group according to gender. d) The correlation analysis between CAPN5, ENPEP, FUT6, GDPD3, GPD1L, HAO2, HNRNPA1, NAGA, and RAB17 protein abundances and MoCA for each group according to the gender. The red-bordered areas show either positive or negative correlations. The unpaired two-samples Wilcoxon test p-values were shown for the comparison between gender and specific proteins. The correlations between potential biomarker expressions with gender, age, disease duration, and MoCA were created with a minimal 0.6 for R^2^ and a maximal 0.05 for p-value calculated using t-distribution with n-2 degrees of freedom as thresholds.

**Supplementary Figure 6.**
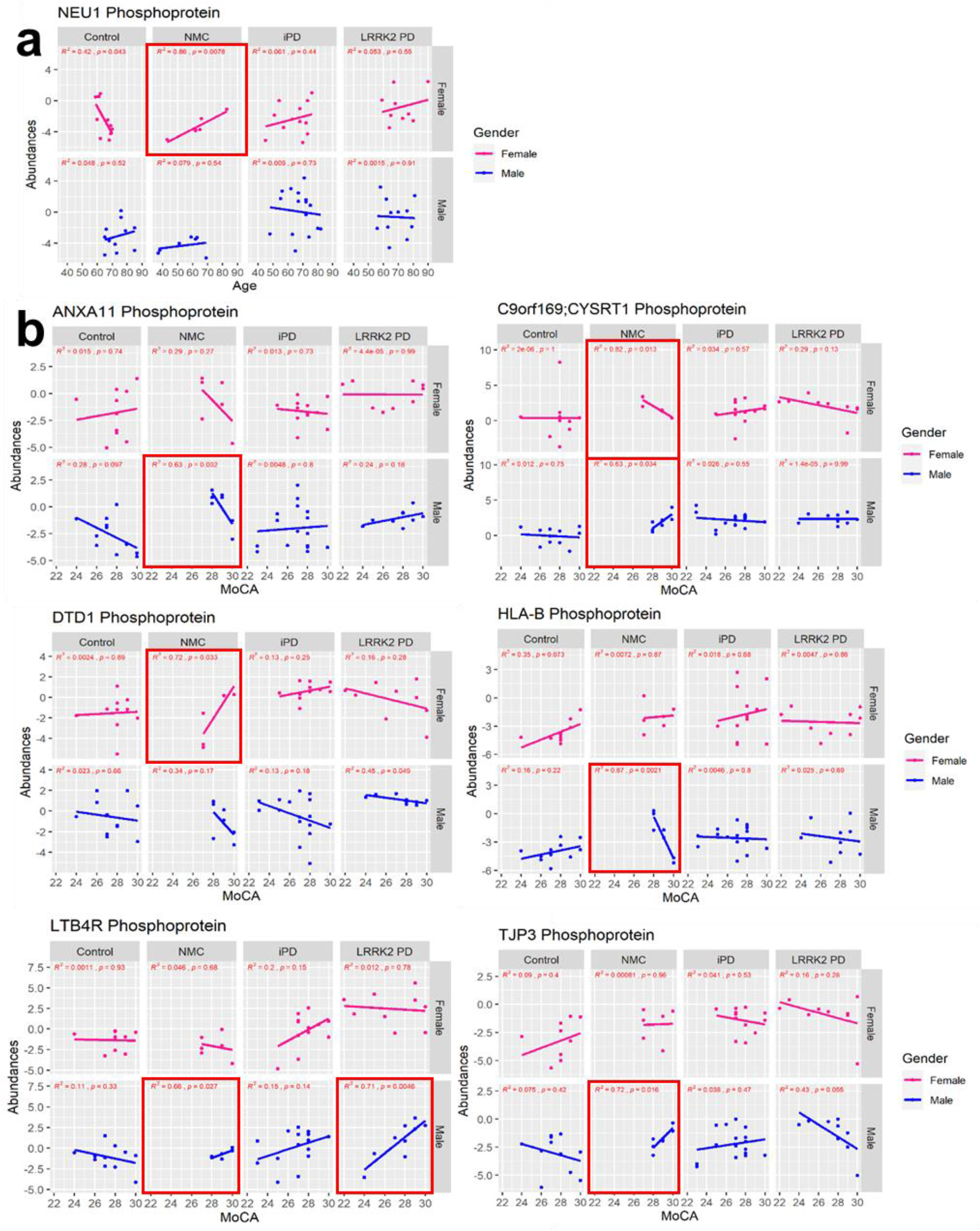
Correlation analysis for select potential phosphoprotein biomarkers from the full data set (training and test sets). a) The correlation analysis between NEU1 phosphoprotein abundances and age for each group according to gender. b) The correlation analysis between ANXA11, CYSRT1, DTD1, HLA-B, LTB4R, and TJP3 phosphoprotein abundances and MoCA for each group according to the gender. The red-bordered areas show either positive or negative correlations. The correlations between potential biomarker expressions with age and MoCA were created with a minimal 0.6 for R^2^ and a maximal 0.05 for p-value calculated using t-distribution with n-2 degrees of freedom as thresholds.

**Supplementary Figure 7.**
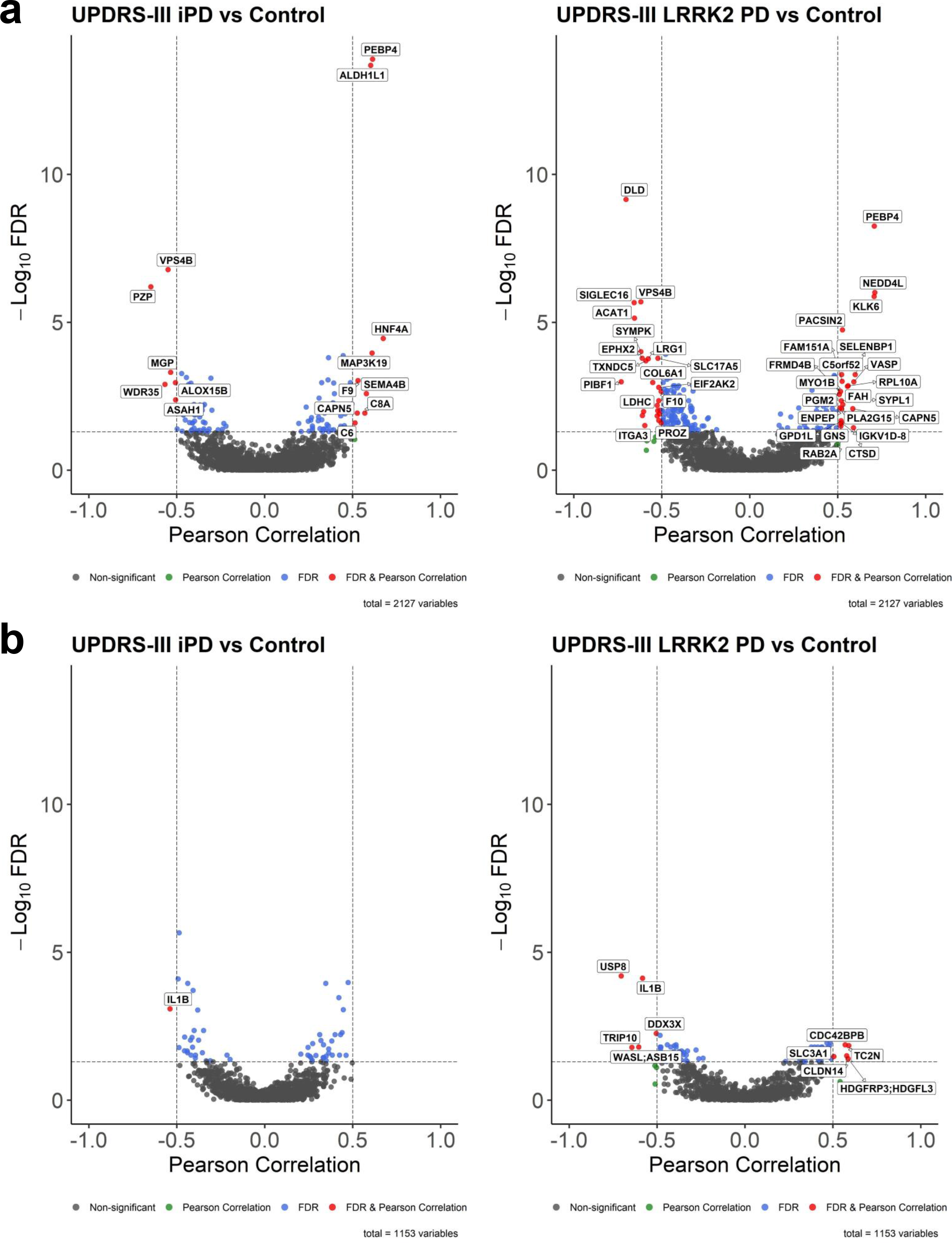
Correlations with clinical parameter, UPDRS-III. Pearson correlation scores and associated *P*-values [-log_10_] of all a) protein and b) phosphoprotein intensities with the UPDRS-III score. Either all iPD patients (left) or LRRK2 PD patients (right) were included. Significantly correlated proteins with an FDR of 5% after Benjamin-Hochberg correction and Pearson correlation more than 0.5 are labeled.

**Supplementary Figure 8.**
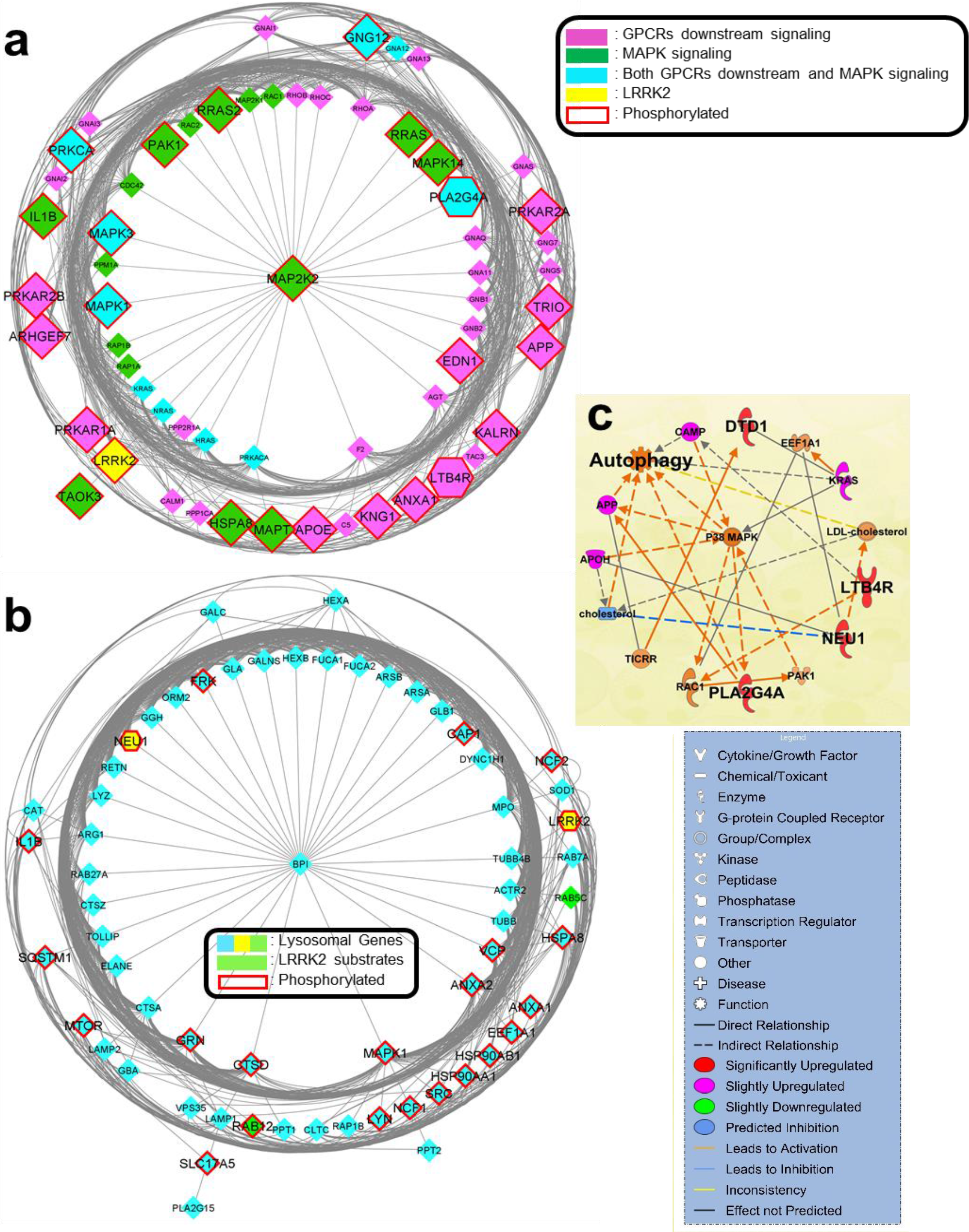
Phosphoprotein disease biomarker network and pathway analyses. Enriched networks include a) GPCRs and MAPK signaling pathways and b) lysosome regulation and lysosomal disorder. c) IPA pathway analysis of the phosphoprotein disease markers related to the autophagy pathway.

**Supplementary Figure 9.**
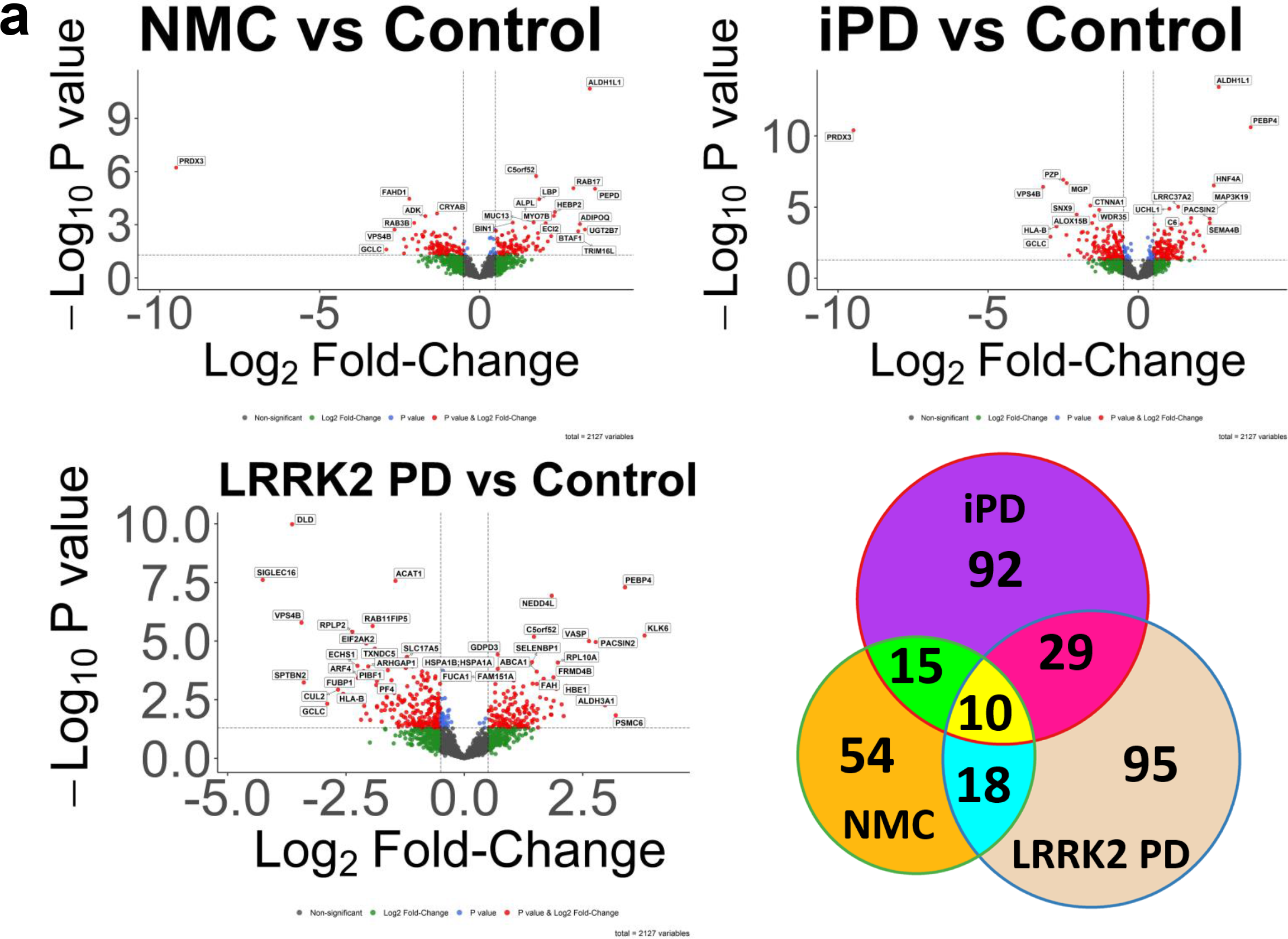
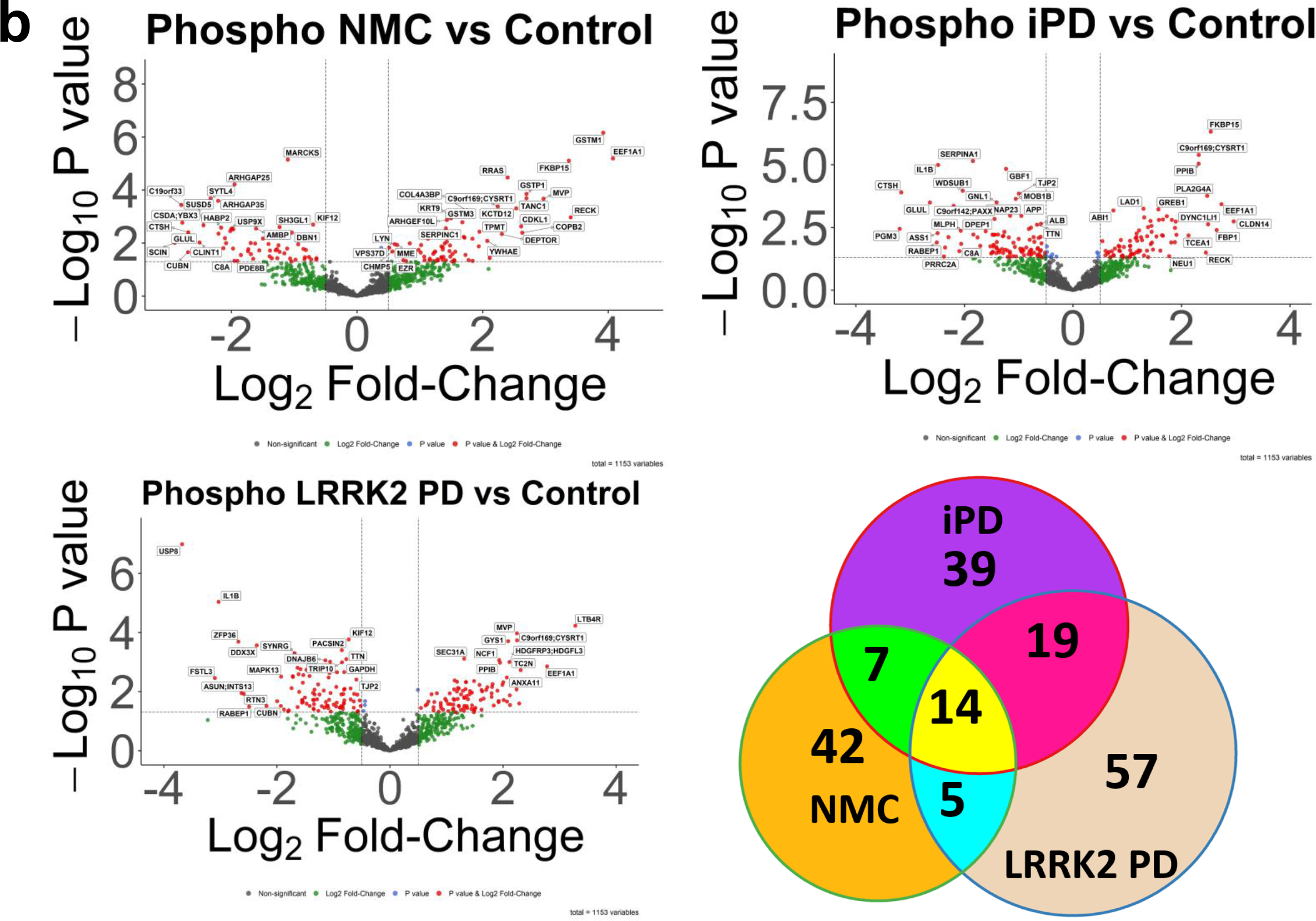
Biosignature study design created on the training set. All three categories: NMC, iPD, and LRRK2 PD were compared to the Control group for (a) proteins and (b) phosphoproteins. Volcano plots were created for each comparison with cut-off values of Student’s two-tailed t-test p-value = 0.05 and log base 2 fold-change = 0.5, which equals to ∼1.414 fold-change. Significantly up-regulated phosphoproteins from the three volcano plots were overlapped in Venn diagrams (see Supplementary Data 14 for overlapping proteins and phosphoproteins).

**Supplementary Figure 10.**
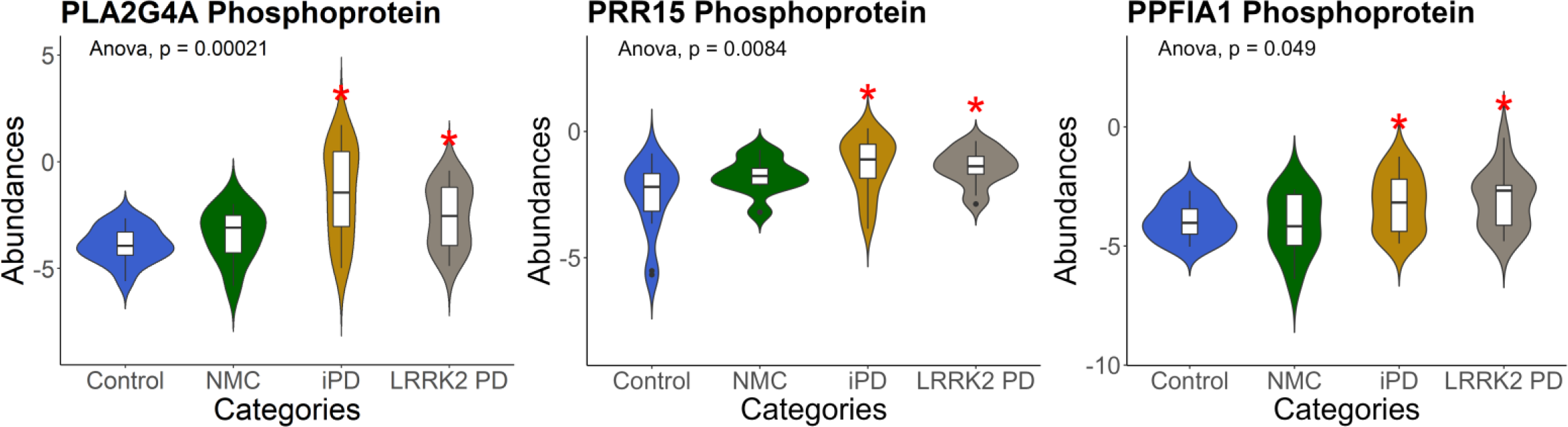
The additional selected top disease biomarkers acquired from the training set. Violin plots of the statistically upregulated proteins and phosphoproteins from the training set in PD regardless of the *LRRK2*-G2019S mutation (disease markers). The unpaired two-samples Wilcoxon test p-values and the one-way ANOVA p-value were included for each of the violin plots.

**Supplementary Figure 11.**
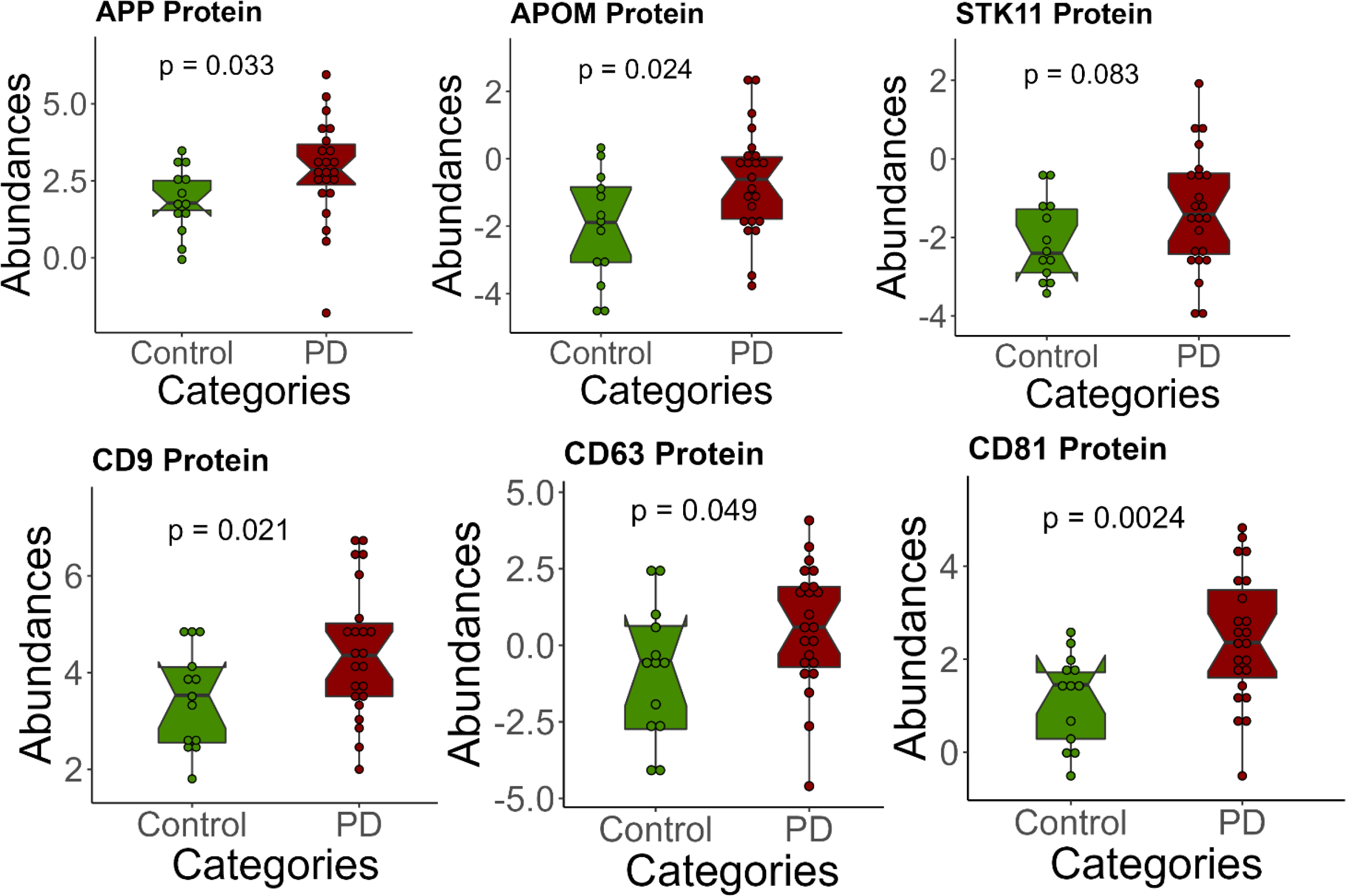
The additional targeted quantitation of potential disease biomarkers. Boxplots of the statistically significant upregulated disease markers (proteins). The Student’s two tailed t-test was used to calculate all P values.

**Supplementary Figure 12.**
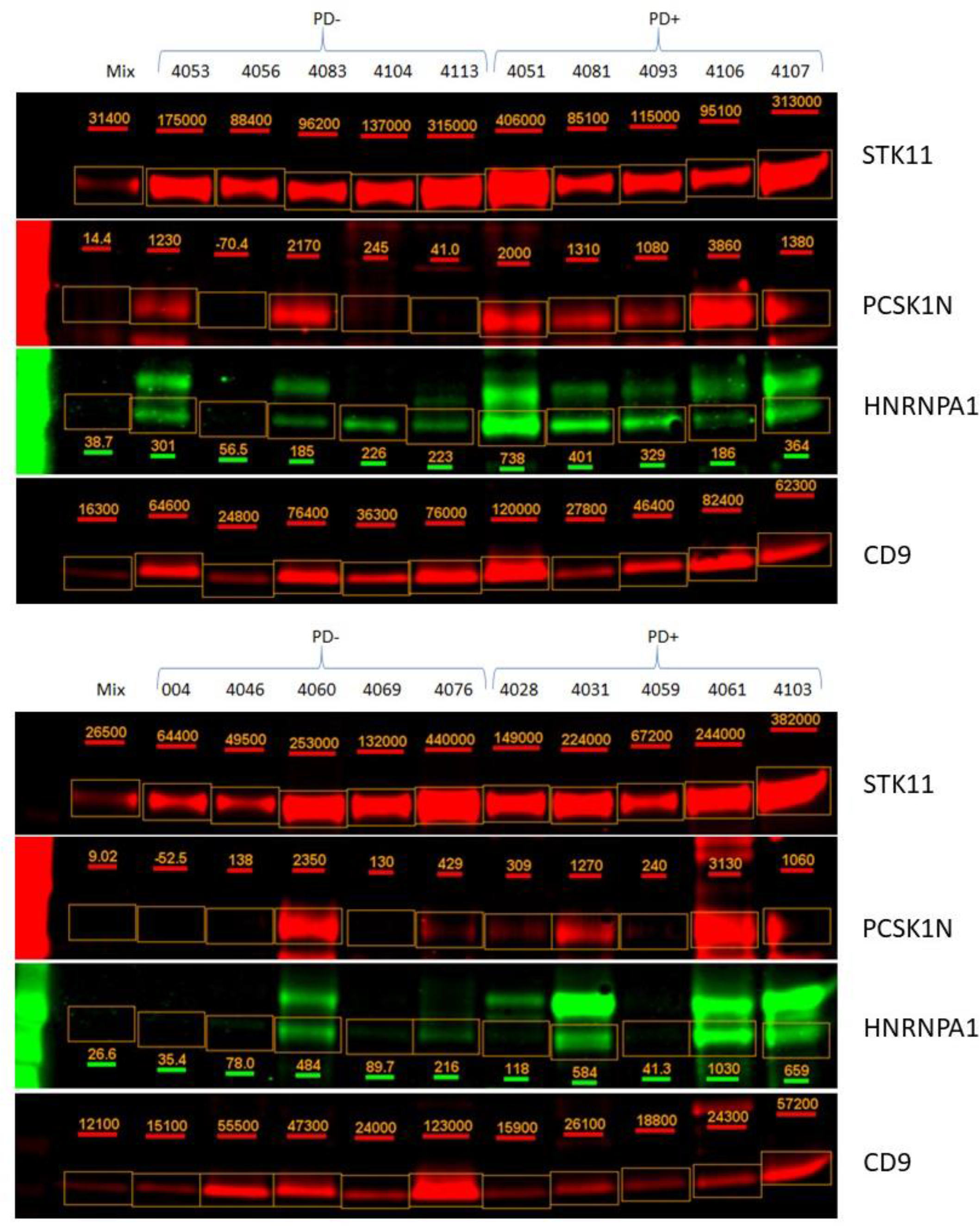
Western blot analysis of CD9, STK11, PCSK1N, and HNRNPA1. All 20 urine EV samples were analyzed by Western Blot with anti-CD9, anti-STK11, anti-PCSK1N, and anti-HNRNPA1 antibodies (2 blots for each type). An equal amount of pooled urine EVs was loaded in lane 1 of each gel.

**Supplementary Figure 13.**
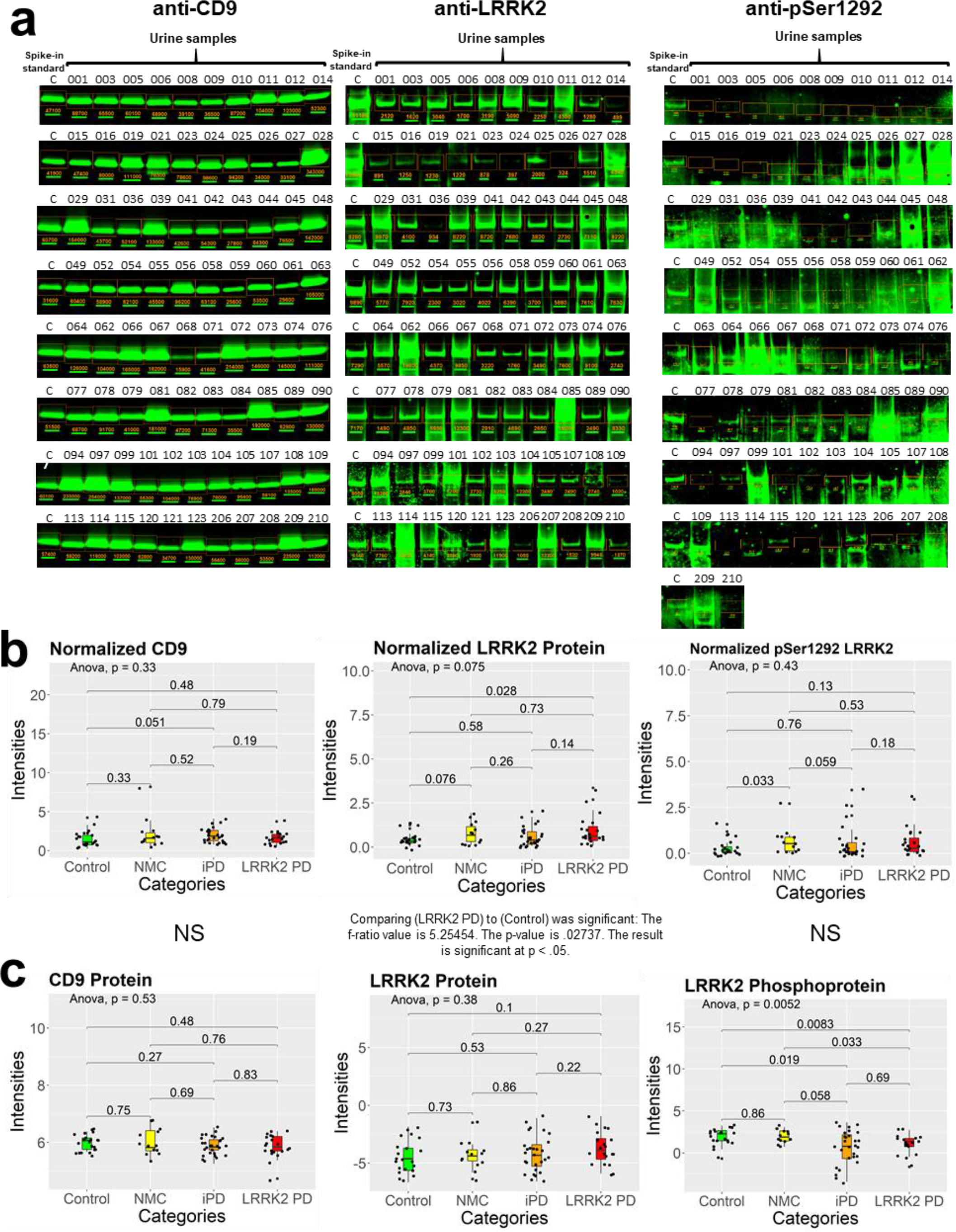
Western blot analysis of CD9, total LRRK2 and pSer1292-LRRK2. All 82 urine EV samples were analyzed by Western Blot with anti-CD9, anti-LRRK2, and anti-pSer1292-LRRK2 antibodies (8 blots for each type). An equal amount of a spike-in standard was loaded in lane 1 of each gel (pooled urine EVs for CD9 blots, recombinant LRRK2 for LRRK2 blots, and autophosphorylated recombinant LRRK2 for pSer1292-LRRK2 blots). a) Western blots from each analyzed target protein. b) Western blot-based quantitative comparison across all samples after normalization with an internal standard (See Supplementary Data 17 for table format). c) Mass spectrometry-based quantitative comparison across all samples after normalization with an internal standard. The Student’s two-tailed t-test p-values and the one-way ANOVA p-value were included on each of the boxplots.

**Supplementary Figure 14.**
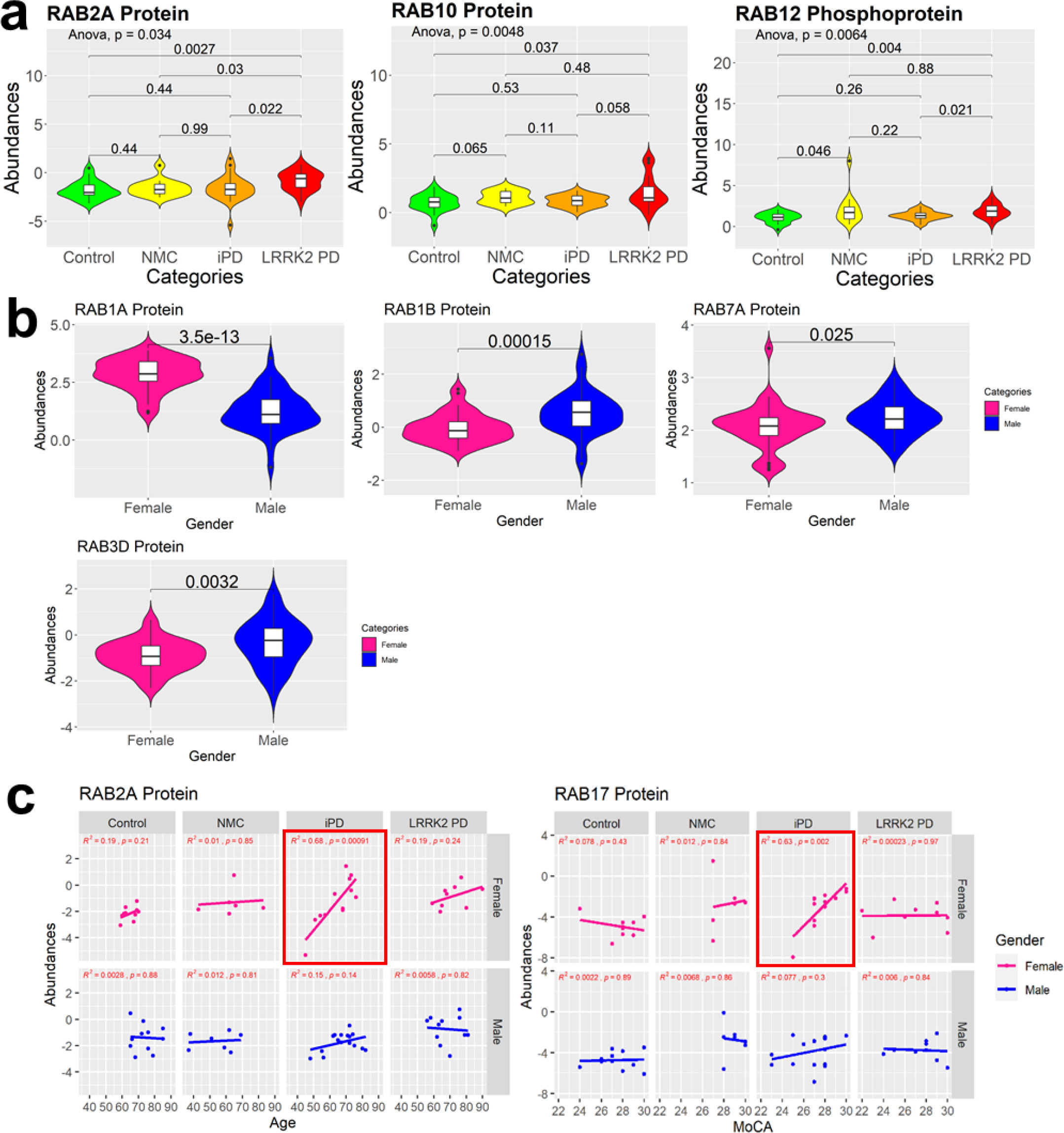
Significantly changing Rab proteins and phosphoproteins. a) The EV levels of Rab2A and Rab10 were significantly higher than the control. b) Rab1A was expressed at lower levels in males; Rab1B, Rab3D, and Rab7A were expressed at higher levels in males. c) The correlation analysis between Rab2A protein abundances and age for each group according to gender. d) The correlation analysis between Rab17 protein abundances and MoCA for each group according to gender. The red-bordered areas show positive correlations. The correlations between potential biomarker expressions with gender, age, and MoCA were created with a minimal 0.6 for R^2^ and a maximal 0.05 for p-value calculated using t-distribution with n-2 degrees of freedom as thresholds.

**Supplementary Table 1.** List of the feature selection inputs (potential biomarkers) for both disease and risk biomarkers. Intermediate results after backward feature elimination and before exhaustive feature selection are colored green. The top biomarkers are bordered with black lines.

| Input Disease Biomarkers |  | Input Risk Biomarkers |
| --- | --- | --- |
| TGM3 | UGP2 | ALDH1L1 |
| ABCA1 | CAPN5 | TRIM16L |
| PCSK1N | GALNT7 | FAM157C |
| HNRNPA1 | FLOT1 | ALPL |
| SLC22A13 | FAM151A | AMN |
| FUT6 | pCLDN14 | ACPP |
| RPS4X | pDYNC1LI1 | UBE2N |
| NCCRP1 | pPLA2G4A | B4GALT3 |
| RALA | pNEU1 | CD5L |
| SAA4 | pLTB4R | ITIH1 |
| STK11 | pEDN1 | HLA-C |
| ITCH | pPRG4 | GNB2 |
| FABP3 | pLAD1 | ARL15 |
| C4BPA | pPRR15 | COL1A2 |
| KLK10 | pSSB | GNG7 |
| UCHL1 | pTMPO | pRECK |
| IGF1 | pTBC1D9B | pFKBP15 |
| C6orf211;ARMT1 | pCLIC6 | pTRPM7 |
| CC2D1A | pTJP3 | pCLEC3B |
| ACAT2 | pDKC1 | pSERPINC1 |
| VASP | pPRKAR2A | pPLEKHO2 |
| IDE | pPPFIA1 | pMICALL1 |
| ERBB2 | pFNBP1 |  |
| PGM2 | pSPPL2B |  |

**Supplementary Table 2.** List of the proteins targeted by PRM-MS.

| Gene Name | Reason |
| --- | --- |
| HNRNPA1 | Top Disease Biomarker |
| PCSK1N | Top Disease Biomarker |
| ALDH1L1 | Top Risk Biomarker |
| IGF1 | Potential Disease Biomarker |
| STK11 | Potential Disease Biomarker |
| ALPL | Potential Risk Biomarker |
| B4GALT3 | Potential Risk Biomarker |
| FN1 | Co-purified with EV |
| A2M | EV Marker |
| ALB | EV Marker |
| CD63 | EV Marker |
| CD81 | EV Marker |
| CD9 | EV Marker |
| MSN | EV Marker |
| APOE | Involved in PD Pathway |
| APOM | Involved in PD Pathway |
| APOA1 | Known PD Biomarker |
| APP | Known PD Biomarker |
| HNF4A | Known PD Biomarker |
| PARK7 | Known PD Biomarker |

**Supplementary Table 3.** List of the 34 identified Rab GTPases, 12 of which are known to be LRRK2 substrates, and eight phosphorylated Rab GTPases.

| Proteins |  |  | Phosphoproteins |  |  |
| --- | --- | --- | --- | --- | --- |
| Accession | Gene Symbol | # PSMs | Accession | Gene Symbol | # PSMs |
| P61026 | RAB10 | 1740 | P61026 | RAB10 | 57 |
| Q15907 | RAB11B | 1400 | Q6IQ22 | RAB12 | 384 |
| Q6IQ22 | RAB12 | 37 | P51153 | RAB13 | 13 |
| P51153 | RAB13 | 171 | Q14966-1 | RAB7L1; RAB29 | 135 |
| P61106 | RAB14 | 612 | Q86YS6 | RAB43 | 3 |
| Q9H0T7 | RAB17 | 26 | P51149 | RAB7A | 2 |
| Q9NP72 | RAB18 | 25 | P61006 | RAB8A | 290 |
| P62820 | RAB1A | 622 | Q92930 | RAB8B | 34 |
| Q9H0U4 | RAB1B | 828 |  |  |  |
| Q9NX57 | RAB20 | 2 |  |  |  |
| Q9UL25 | RAB21 | 166 |  |  |  |
| Q9UL26 | RAB22A | 55 |  |  |  |
| Q9ULC3 | RAB23 | 24 |  |  |  |
| P57735 | RAB25 | 107 |  |  |  |
| P51159-1 | RAB27A | 271 |  |  |  |
| O00194 | RAB27B | 159 |  |  |  |
| P61019-1 | RAB2A | 273 |  |  |  |
| Q13636 | RAB31 | 3 |  |  |  |
| Q13637 | RAB32 | 8 |  |  |  |
| Q9BZG1 | RAB34 | 5 |  |  |  |
| Q15286 | RAB35 | 437 |  |  |  |
| P20337 | RAB3B | 127 |  |  |  |
| O95716 | RAB3D | 331 |  |  |  |
| Q86YS6 | RAB43 | 38 |  |  |  |
| P20338 | RAB4A | 38 |  |  |  |
| P20339 | RAB5A | 433 |  |  |  |
| P61020 | RAB5B | 564 |  |  |  |
| P51148-2 | RAB5C | 1444 |  |  |  |
| P20340-2 | RAB6A | 58 |  |  |  |
| P51149 | RAB7A | 1861 |  |  |  |
| O14966-1 | RAB7L1; RAB29 | 268 |  |  |  |
| P61006 | RAB8A | 1016 |  |  |  |
| Q92930 | RAB8B | 576 |  |  |  |
| P51151 | RAB9A | 24 |  |  |  |

LRRK2 Substrates Quantified

## References

1. Zeng, X. S., Geng, W. S., Jia, J. J., Chen, L. & Zhang, P. P. Cellular and molecular basis of neurodegeneration in Parkinson disease. Front. Aging Neurosci. 10, 1–16 (2018).

2. Rui, Q., Ni, H., Li, D., Gao, R. & Chen, G. The Role of LRRK2 in Neurodegeneration of Parkinson Disease. Curr. Neuropharmacol. 16, 1348–1357 (2018).

3. Houlden, H. & Singleton, A. B. The genetics and neuropathology of Parkinson’s disease. Acta Neuropathol. 124, 325–338 (2012).

4. Adams, B. et al. Parkinson’s disease: A systemic inflammatory disease accompanied by bacterial inflammagens. Front. Aging Neurosci. 10, 1–17 (2019).

5. Tibar, H. et al. Non-motor symptoms of Parkinson’s Disease and their impact on quality of life in a cohort of Moroccan patients. Front. Neurol. 9, 1–12 (2018).

6. Zhong, J. et al. Integrated profiling of single cell epigenomic and transcriptomic landscape of Parkinson’s disease mouse brain. bioRvix (2020) doi:10.1101/2020.02.04.933259.

7. Reeve, A., Simcox, E. & Turnbull, D. Ageing and Parkinson’s disease: Why is advancing age the biggest risk factor? Ageing Res. Rev. 14, 19–30 (2014).

8. Hughes, R. C. Parkinson’s Disease and its Management. Bmj 308, 281 (1994).

9. Marras, C. et al. Prevalence of Parkinson’s disease across North America. *npj Park*. Dis. 4, 1–7 (2018).

10. Burbulla, L. F. & Krüger, R. Converging environmental and genetic pathways in the pathogenesis of Parkinson’s disease. J. Neurol. Sci. 306, 1–8 (2011).

11. Chang, D. et al. A meta-analysis of genome-wide association studies identifies 17 new Parkinson’s disease risk loci. Nat. Genet. 49, 1511–1516 (2017).

12. Satake, W. et al. Genome-wide association study identifies common variants at four loci as genetic risk factors for Parkinson’s disease. Nat. Genet. 41, 1303–1307 (2009).

13. Simón-Sánchez, J. et al. Genome-wide association study reveals genetic risk underlying Parkinson’s disease. Nat. Genet. 41, 1308–1312 (2009).

14. Price, A., Manzoni, C., Cookson, M. R. & Lewis, P. A. The LRRK2 signalling system. Cell Tissue Res. 373, 39–50 (2018).

15. Marín, I., Egmond, W. N. & Haastert, P. J. M. The Roco protein family: a functional perspective. FASEB J. 22, 3103–3110 (2008).

16. Alessi, D. R. & Sammler, E. LRRK2 kinase in Parkinson’s disease. Science (80-.). 360, 36–37 (2018).

17. Migheli, R. et al. LRRK2 Affects Vesicle Trafficking, Neurotransmitter Extracellular Level and Membrane Receptor Localization. PLoS One 8, e77198 (2013).

18. Margolis, L. & Sadovsky, Y. The biology of extracellular vesicles: The known unknowns. PLOS Biol. 17, 1–12 (2019).

19. Zhang, Y., Wu, X. & Tao, W. A. Characterization and applications of extracellular vesicle proteome with post-translational modifications. TrAC - Trends Anal. Chem. 107, 21–30 (2018).

20. Santucci, L. et al. Biological surface properties in extracellular vesicles and their effect on cargo proteins. Sci. Rep. 9, 1–12 (2019).

21. Yang, K. S. et al. Multiparametric plasma EV profiling facilitates diagnosis of pancreatic malignancy. Sci. Transl. Med. 9, eaal3226 (2017).

22. Verma, M., Lam, T. K., Hebert, E. & Divi, R. L. Extracellular vesicles: Potential applications in cancer diagnosis, prognosis, and epidemiology. BMC Clin. Pathol. 15, 1–9 (2015).

23. Xu, R., Greening, D. W., Zhu, H.-J., Takahashi, N. & Simpson, R. J. Extracellular vesicle isolation and characterization: toward clinical application. J. Clin. Invest. 126, 1152–1162 (2016).

24. Lin, J. et al. Exosomes: Novel Biomarkers for Clinical Diagnosis. Sci. World J. 2015, (2015).

25. Bouhaddou, M. et al. The Global Phosphorylation Landscape of SARS-CoV-2 Infection. Cell 182, 685–712.e19 (2020).

26. Chen, I. H. et al. Phosphoproteins in extracellular vesicles as candidate markers for breast cancer. Proc. Natl. Acad. Sci. U. S. A. 114, 3175–3180 (2017).

27. Iliuk, A. et al. Plasma-Derived Extracellular Vesicle Phosphoproteomics through Chemical Affinity Purification. J. Proteome Res. (2020) doi:10.1021/acs.jproteome.0c00151.

28. Hadisurya, M. et al. Data-independent acquisition phosphoproteomics of urinary extracellular vesicles enables renal cell carcinoma grade differentiation. medRxiv 2022.08.15.22278799 (2022) doi:10.1101/2022.08.15.22278799.

29. Stuendl, A. et al. Induction of α-synuclein aggregate formation by CSF exosomes from patients with Parkinson’s disease and dementia with Lewy bodies. Brain 139, 481–494 (2016).

30. Cao, Z. et al. α-Synuclein in salivary extracellular vesicles as a potential biomarker of Parkinson’s disease. Neurosci. Lett. 696, 114–120 (2019).

31. Jiang, C. et al. Serum neuronal exosomes predict and differentiate Parkinson’s disease from atypical parkinsonism. J. Neurol. Neurosurg. Psychiatry 91, 720–729 (2020).

32. Alcalay, R. N. et al. Higher Urine bis(Monoacylglycerol)Phosphate Levels in LRRK2 G2019S Mutation Carriers: Implications for Therapeutic Development. Mov. Disord. 35, 134–141 (2020).

33. Decramer, S. et al. Urine in clinical proteomics. Mol. Cell. Proteomics 7, 1850–1862 (2008).

34. An, M. & Gao, Y. Urinary Biomarkers of Brain Diseases. Genomics, Proteomics Bioinforma. 13, 345–354 (2015).

35. Wang, S., Kojima, K., Mobley, J. A. & West, A. B. Proteomic analysis of urinary extracellular vesicles reveal biomarkers for neurologic disease. EBioMedicine 45, 351– 361 (2019).

36. Upadhya, R. & Shetty, A. K. Extracellular vesicles for the diagnosis and treatment of Parkinson’s disease. Aging Dis. 12, 1438–1450 (2021).

37. Wu, X., Li, L., Iliuk, A. & Tao, W. A. Highly Efficient Phosphoproteome Capture and Analysis from Urinary Extracellular Vesicles. J. Proteome Res. 17, 3308–3316 (2018).

38. Buljan, M., Blattmann, P., Aebersold, R. & Boutros, M. Systematic characterization of pan-cancer mutation clusters. Mol. Syst. Biol. 14, 1–19 (2018).

39. Le Large, T. Y. S. et al. Key biological processes driving metastatic spread of pancreatic cancer as identified by multi-omics studies. Semin. Cancer Biol. 44, 153–169 (2017).

40. Pishvaian, M. J. et al. Molecular profiling of patients with pancreatic cancer: Initial results from the know your tumor initiative. Clin. Cancer Res. 24, 5018–5027 (2018).

41. Chen, F., Chandrashekar, D. S., Varambally, S. & Creighton, C. J. Pan-cancer molecular subtypes revealed by mass-spectrometry-based proteomic characterization of more than 500 human cancers. Nat. Commun. 10, 1–15 (2019).

42. Chen, F. et al. Multiplatform-based molecular subtypes of non-small-cell lung cancer. Oncogene 36, 1384–1393 (2017).

43. Li, J. et al. Characterization of Human Cancer Cell Lines by Reverse-phase Protein Arrays. Cancer Cell 31, 225–239 (2017).

44. Mundt, F. et al. Mass spectrometry–based proteomics reveals potential roles of NEK9 and MAP2K4 in resistance to PI3K inhibition in triple-negative breast cancers. Cancer Res. 78, 2732–2746 (2018).

45. Wulfkuhle, J. D. et al. Molecular analysis of HER2 signaling in human breast cancer by functional protein pathway activation mapping. Clin. Cancer Res. 18, 6426–6435 (2012).

46. Wulfkuhle, J. D. et al. Evaluation of the HER/PI3K/AKT Family Signaling Network as a Predictive Biomarker of Pathologic Complete Response for Patients With Breast Cancer Treated With Neratinib in the I-SPY 2 TRIAL. *JCO Precis*. Oncol. 1–20 (2018) doi:10.1200/po.18.00024.

47. Zagorac, I. et al. In vivo phosphoproteomics reveals kinase activity profiles that predict treatment outcome in triple-negative breast cancer. Nat. Commun. 9, (2018).

48. Huang, K. L. et al. Proteogenomic integration reveals therapeutic targets in breast cancer xenografts. Nat. Commun. 8, (2017).

49. Wang, S. et al. Elevated LRRK2 autophosphorylation in brain-derived and peripheral exosomes in LRRK2 mutation carriers. Acta Neuropathol. Commun. 5, 86 (2017).

50. Fraser, K. B., Moehle, M. S., Alcalay, R. N. & West, A. B. Urinary LRRK2 phosphorylation predicts parkinsonian phenotypes in G2019S LRRK2 carriers. Neurology 86, 994–999 (2016).

51. Fraser, K. B., et al. Ser(P)-1292 LRRK2 in urinary exosomes is elevated in idiopathic Parkinson’s disease. Mov. Disord. 31, 1543–1550 (2016).

52. Virreira Winter, S., et al. Urinary proteome profiling for stratifying patients with familial Parkinson’s disease. EMBO Mol. Med. 2020.08.09.243584 (2021) doi:10.15252/emmm.202013257.

53. Alcalay, R. N. et al. Parkinson disease phenotype in Ashkenazi jews with and without LRRK2 G2019S mutations. Mov. Disord. 28, 1966–1971 (2013).

54. Keerthikumar, S. et al. ExoCarta: A Web-Based Compendium of Exosomal Cargo. J. Mol. Biol. 428, 688–692 (2016).

55. Théry, C., et al. Minimal information for studies of extracellular vesicles 2018 (MISEV2018): a position statement of the International Society for Extracellular Vesicles and update of the MISEV2014 guidelines. J. Extracell. Vesicles 7, (2018).

56. Zhao, M. et al. A comprehensive analysis and annotation of human normal urinary proteome. Sci. Rep. 7, 1–13 (2017).

57. Uhlen, M. et al. Tissue-based map of the human proteome. Science (80-.). 347, 1260419–1260419 (2015).

58. Zhu, Q. et al. The genetic source tracking of human urinary exosomes. Proc. Natl. Acad. Sci. U. S. A. 118, 10–12 (2021).

59. Szklarczyk, D. et al. STRING v11: Protein-protein association networks with increased coverage, supporting functional discovery in genome-wide experimental datasets. Nucleic Acids Res. 47, D607–D613 (2019).

60. Videira, P. A. Q. & Castro-Caldas, M. Linking glycation and glycosylation with inflammation and mitochondrial dysfunction in Parkinson’s disease. Front. Neurosci. 12, 1–20 (2018).

61. Trezzi, J. P. et al. Distinct metabolomic signature in cerebrospinal fluid in early parkinson’s disease. Mov. Disord. 32, 1401–1408 (2017).

62. Everse, J., Liu, C. J. J. & Coates, P. W. Physical and catalytic properties of a peroxidase derived from cytochrome c. Biochim. Biophys. Acta - Mol. Basis Dis. 1812, 1138–1145 (2011).

63. Loeffler, D. A., Camp, D. M. & Conant, S. B. Complement activation in the Parkinson’s disease substantia nigra: An immunocytochemical study. J. Neuroinflammation 3, 1–8 (2006).

64. Sasaki, M. et al. Neuromelanin magnetic resonance imaging of locus ceruleus and substantia nigra in Parkinson’s disease. Neuroreport 17, 1215–1218 (2006).

65. Song, J. & Kim, J. Degeneration of dopaminergic neurons due to metabolic alterations and Parkinson’s disease. Front. Aging Neurosci. 8, 1–11 (2016).

66. Seol, W., Nam, D. & Son, I. Rab GTPases as physiological substrates of LRRK2 kinase. Exp. Neurobiol. 28, 134–145 (2019).

67. Gardet, A. et al. LRRK2 Is Involved in the IFN-γ Response and Host Response to Pathogens. J. Immunol. 185, 5577–5585 (2010).

68. Hakimi, M. et al. Parkinson’s disease-linked LRRK2 is expressed in circulating and tissue immune cells and upregulated following recognition of microbial structures. J. Neural Transm. 118, 795–808 (2011).

69. Cerri, S., Mus, L. & Blandini, F. Parkinson’s Disease in Women and Men: What’s the Difference? J. Parkinsons. Dis. 9, 501–515 (2019).

70. Baldereschi, M. et al. Parkinson’s disease and parkinsonism in a longitudinal study: Two-fold higher incidence in men. Neurology 55, 1358–1363 (2000).

71. Vásquez, K. A., Valverde, E. M., Aguilar, D. V. & Gabarain, H. J. H. Montreal cognitive assessment scale in patients with parkinson disease with normal scores in the mini-mental state examination. Dement. e Neuropsychol. 13, 78–81 (2019).

72. Sun, Y., Vashisht, A. A., Tchieu, J., Wohlschlegel, J. A. & Dreier, L. Voltage-dependent anion channels (VDACs) recruit parkin to defective mitochondria to promote mitochondrial autophagy. J. Biol. Chem. 287, 40652–40660 (2012).

73. Klein, A. D. & Mazzulli, J. R. Is Parkinson’s disease a lysosomal disorder? Brain 141, 2255–2262 (2018).

74. Sarkar, C. et al. PLA2G4A/cPLA2-mediated lysosomal membrane damage leads to inhibition of autophagy and neurodegeneration after brain trauma. Autophagy 16, 466– 485 (2019).

75. Mudannayake, J. M., Mouravlev, A., Fong, D. M. & Young, D. Transcriptional activity of novel ALDH1L1 promoters in the rat brain following AAV vector-mediated gene transfer. Mol. Ther. -Methods Clin. Dev. 3, 16075 (2016).

76. Romano, R. & Bucci, C. Role of EGFR in the Nervous System. Cells 9, (2020).

77. Cassani, E. et al. Cardiometabolic factors and disease duration in patients with Parkinson’s disease. Nutrition 29, 1331–1335 (2013).

78. Klemann, C. J. H. M. et al. Integrated molecular landscape of Parkinson’s disease. *npj Park*. Dis. 3, (2017).

79. He, R. et al. Recent advances in biomarkers for Parkinson’s disease. Front. Aging Neurosci. 10, 1–19 (2018).

80. Carecchio, M. & Comi, C. The role of osteopontin in neurodegenerative diseases. J. Alzheimer’s Dis. 25, 179–185 (2011).

81. Trevor Hastie, Robert Tibshirani, J. F. The Elements of Statistical Learning: Data Mining, Inference, and Prediction. in 57–60 (Springer, 2009).

82. Safari, S., Baratloo, A., Elfil, M. & Negida, A. Evidence Based Emergency Medicine; Part 5 Receiver Operating Curve and Area under the Curve. Emerg. (Tehran, Iran) 4, 111–3 (2016).

83. Santiago, J. A. & Potashkin, J. A. Network-based metaanalysis identifies HNF4A and PTBP1 as longitudinally dynamic biomarkers for Parkinson’s disease. Proc. Natl. Acad. Sci. U. S. A. 112, 2257–2262 (2015).

84. Vacchi, E. et al. Immune profiling of plasma-derived extracellular vesicles identifies Parkinson disease. Neurol. Neuroimmunol. neuroinflammation 7, (2020).

85. Di Maio, R. et al. LRRK2 activation in idiopathic Parkinson’s disease. Sci. Transl. Med. 10, 1–13 (2018).

86. Petridi, S. et al. Neurodegeneration caused by LRRK2-G2019S requires Rab10 in select dopaminergic neurons. bioRxiv 586073 (2019) doi:10.1101/586073.

87. Cova, I. & Priori, A. Diagnostic biomarkers for Parkinson’s disease at a glance: where are we? J. Neural Transm. 125, 1417–1432 (2018).

88. Oberg, A. L. & Vitek, O. Statistical design of quantitative mass spectrometry-based proteomic experiments. J. Proteome Res. 8, 2144–2156 (2009).

89. Zeringer, E. Methods for the extraction and RNA profiling of exosomes. World J. Methodol. 3, 11 (2013).

90. Rauniyar, N. Parallel reaction monitoring: A targeted experiment performed using high resolution and high mass accuracy mass spectrometry. Int. J. Mol. Sci. 16, 28566–28581 (2015).

91. Deutsch, E. W., Lam, H. & Aebersold, R. PeptideAtlas: A resource for target selection for emerging targeted proteomics workflows. EMBO Rep. 9, 429–434 (2008).

92. MacLean, B., et al. Skyline: An open source document editor for creating and analyzing targeted proteomics experiments. Bioinformatics 26, 966–968 (2010).

93. Tyanova, S. et al. The Perseus computational platform for comprehensive analysis of (prote)omics data. Nat. Methods 13, 731–740 (2016).

94. R Core Team. R: A Language and Environment for Statistical Computing. (2013).

95. Hadley Wickham. ggplot2: Elegant Graphics for Data Analysis. (Springer-Verlag New York, 2016).

96. Kassambara, A. ggpubr: ‘ggplot2’ Based Publication Ready Plots. (2020).

97. Blighe, K. EnhancedVolcano: Publication-ready volcano plots with enhanced colouring and labeling. (2018).

98. Robin, X. et al. pROC: an open-source package for R and S+ to analyze and compare ROC curves. BMC Bioinformatics 12, 77 (2011).

99. Jonathan Swinton. Vennerable: Venn and Euler area-proportional diagrams. (2019).

100. Zuguang Gu, Lei Gu, Roland Eils, Matthias Schlesner, B. B. circlize implements and enhances circular visualization in R. Bioinformatics 30, 2811–2812 (2014).

101. Shannon, P. Cytoscape: A Software Environment for Integrated Models of Biomolecular Interaction Networks. Genome Res. 13, 2498–2504 (2003).

102. Krämer, A., Green, J., Pollard, J. & Tugendreich, S. Causal analysis approaches in ingenuity pathway analysis. Bioinformatics 30, 523–530 (2014).

103. Van Rossum, G. & Drake Jr, F. L. Python tutorial. Cent. voor Wiskd. en Inform. Amsterdam, Netherlands (1995).

104. Anaconda Software Distribution. Anaconda Documentation https://docs.anaconda.com/ (2020).

105. Kluyver, T., et al. Jupyter Notebooks -- a publishing format for reproducible computational workflows. in Positioning and Power in Academic Publishing: Players, Agents and Agendas (ed. Schmidt, F. L. and B.) 87–90 (IOS Press, 2016).

106. McKinney, W. Data Structures for Statistical Computing in Python. in 56–61 (2010). doi:10.25080/Majora-92bf1922-00a.

107. Harris, C. R. et al. Array programming with NumPy. Nature 585, 357–362 (2020).

108. Hunter, J. D. Matplotlib: A 2D Graphics Environment. Comput. Sci. Eng. 9, 90–95 (2007).

109. Inc., P. T. Collaborative data science. https://plot.ly (2015).

110. Pedregosa, F. et al. Scikit-learn: Machine learning in Python. J. Mach. Learn. Res. 12, 2825--2830 (2011).

111. Raschka, S. MLxtend: Providing machine learning and data science utilities and extensions to Python’s scientific computing stack. J. Open Source Softw. 3, 638 (2018).

112. Chen, T. & Guestrin, C. XGBoost: A Scalable Tree Boosting System. in Proceedings of the 22nd ACM SIGKDD International Conference on Knowledge Discovery and Data Mining vol. 42 785–794 (ACM, 2016).

